# ProtoDrift: Evaluating the impact of small-to-big adjustments to chemotherapy protocols

**DOI:** 10.1101/2025.05.09.25327311

**Authors:** Alice Rogier, Elisabeth Ashton, Corine Barat, Romain Griffier, Vianney Jouhet, Elena Paillaud, Brigitte Sabatier, Julien Taïeb, Éric Zapletal, Bastien Rance, Adrien Coulet

## Abstract

Adjustments to chemotherapy protocols are common to adapt treatments to individual patient needs, yet consensus on the impact of such adjustements is lacking. We introduce ProtoDrift, a novel metric that quantifies time and dose adjustments in chemotherapies. This weighted distance enables an assessment of their impact on patient outcomes, offering more detailed analyses than the traditional Relative Dose Intensity (RDI). We compared ProtoDrift and RDI prediction performances through survival analyses on 20,808 patients across 38 groups, categorised by cancer location and treatment line at two hospitals. Without optimisation, ProtoDrift achieves either comparable or better prediction results in 71% of patient groups (27 out of 38). Once optimised, ProtoDrift surpasses the RDI C-index predictions in 89% (16 out of 18) of patient groups from the first hospital. This study confirms ProtoDrift as an advanced tool for refining chemotherapy regimen design, highlighting the critical role of time adjustments in patient outcomes.

## 1 Introduction

Chemotherapy aims at finding the delicate balance between tumour reduction and minimised side effects through the use of cytotoxic drugs. A common strategy to achieve this trade-off is to combine various drugs in a timely manner.^1^ Indeed, chemotherapy regimens are precisely designed to specify the molecules, their dosages, modes and timing of administration in the form of cycles of treatment, aligned with cancer cell life cycles.^2,3^ While regimens are usually defined by their prototypical cycle, the overall chemotherapy protocol consists in the repetition of this cycle over time.

Despite the fact that protocols are precisely defined, their implementation in patient treatment often undergo modifications, which arise for various reasons such as toxicity, accommodations for patient convenience, or operational constraints within healthcare facilities.^4,5,6^ Figure 1 depicts an example of chemotherapy protocol and how its implementation in a patient may drift over time for various reasons.

**Figure 1:**
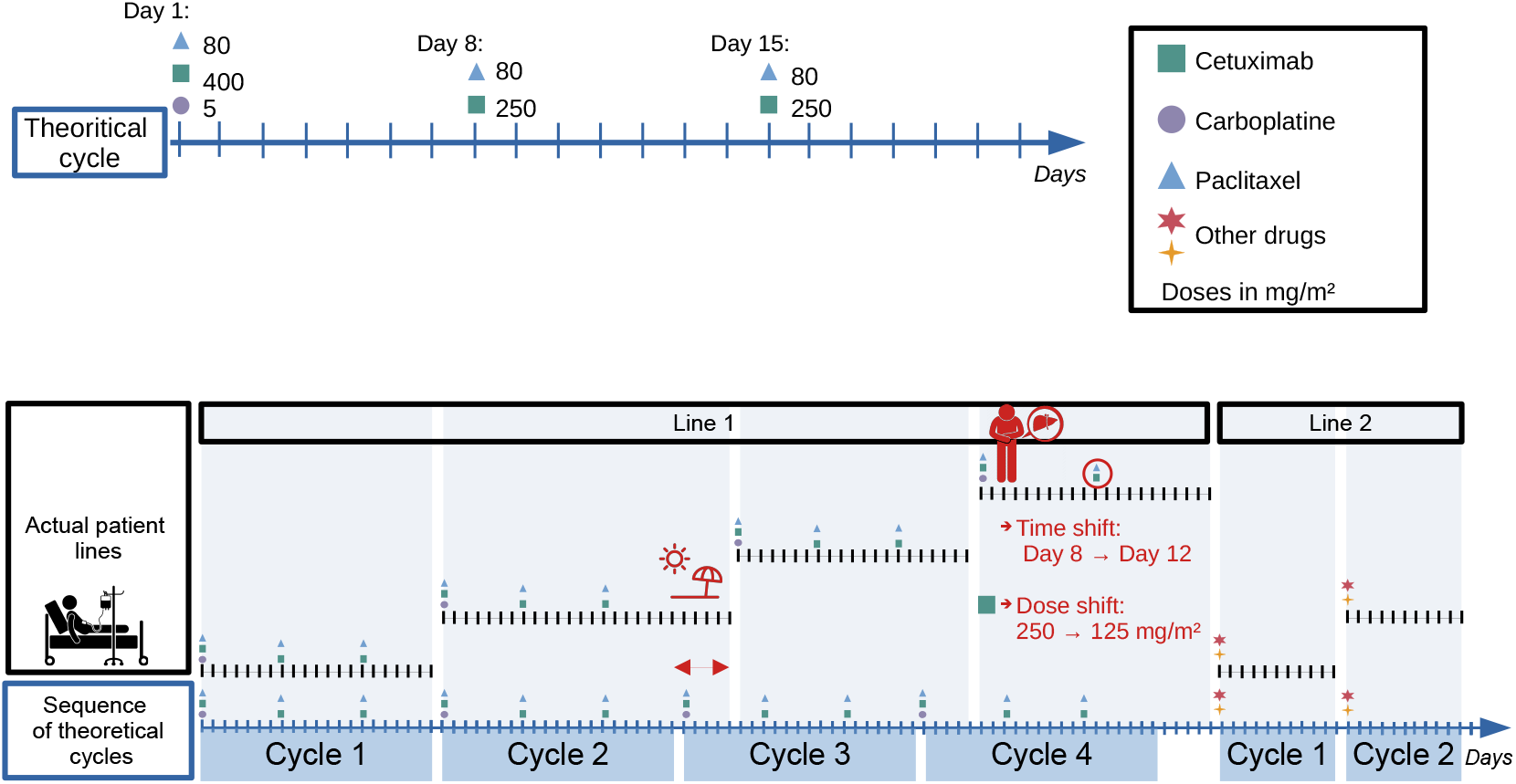
(Top) Graphical description of the regimen 1465 used to treat head and neck cancer. It is 21 days long and composed of various administrations including Cetuximab at day 1 (400mg/m^2^) day 8 (250), day 15 (250) and Paclitaxel at day 1, day 8, day 15 (80mg/m^2^). (Bottom) Comparison between lines (Line 1 and Line 2) followed by a fictive patient and the protocol (*i*.*e*. the sequence of theoretical cycles defined by the regimen itself). In this scenario, cycle 3 is delayed because the patient is in family holidays. In cycle 4, the oncologist decides to change the day of Cetuximab and Paclitaxel administrations, and to reduce the dose of Cetuximab, due to an hepatotoxicity. Both drugs are administrated only twice, instead of three times during this cycle. At the end of cycle 4, after noticing that the tumour is not shrinking fast enough, the medical expert team decides to change the regimen. Thus, a new line of treatment (Line 2) is started.

Adjustment to the protocols are also motivated by empirical hypotheses and vary depending on cancer locations, patient demographics, and other factors. For instance, research on early breast cancer suggests that reducing intervals between treatment cycles can improve patient outcomes.^7^ Meanwhile, other studies have explored how dose reductions for specific cancer treatments can maintain efficacy, while reducing costs, illustrating the range of strategies that might be employed.^8^ Within the diversity of possible adjustments, some are well evaluated but others that are based on clinical experience or habits might lack of consensus.^9^ This highlights the necessity for tools capable of providing fine-grained description of the various types of adjustments and thus enabling the evaluation of their impact on the clinical evolution of patients.

The Relative Dose Intensity (RDI) is the current and standard way to assess the adherence to chemotherapy protocols.^10,11,12^ RDI relies on the dose intensity, denoted DI, that is defined for a given anti-cancer drug *d* as the total dose delivered on a defined period of time in weeks since the start of the treatment line.^13^ In other terms:

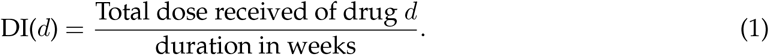

The RDI itself is the ratio of the DI received by a patient on the planned DI, *i*.*e*., the DI that would have been received in the case of a full adherence to the protocol, or

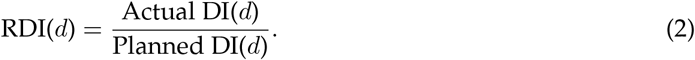

The All Drugs Relative Dose Intensity (ADRDI) is defined for a protocol as the arithmetic mean of the RDIs of the anti-cancer drugs administrated in this protocol:

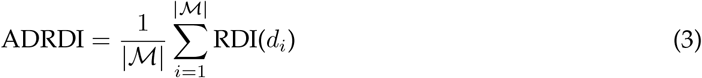

with *ℳ* the set of administrated drugs. Many studies employ the RDI, in combination with a threshold, usually set to 80%, to distinguish adherent from non-adherent patients in survival analyses.^14,15,16,17^ However, this RDI approach oversimplifies the complex dynamics of chemotherapy treatment, focusing on dosage without fully accounting for the timing of administrations and its potential impacts on patient outcomes.

We propose in this article ProtoDrift, a more nuanced approach to quantify treatment deviations from the planned treatment, assigning weighted dissimilarities to each type of variance, aiming at small-to-large variations from the protocol, which could have been motivated by medical care. The finding of the optimal weights offers a detailed view of the relative importance of the various type of adjustments made during chemotherapy. In this sense ProtoDrift may enhance our understanding of treatment responses and guide more effective patient management.

## 2 Results

### 2.1 Patient group characteristics

We computed ADRDI and Naive ProtoDrift (NP) (ProtoDrift without optimised weights) across 38 patient groups from two French University hospitals, the European Hospital Georges Pompidou (HEGP) and the University Hospital of Bordeaux (UHB). Patients groups are formed based on tumour location and treatment line number. The study is composed of 18 patient groups from HEGP hospital, and 20 patients groups from UHB (see Data sources and study design Section in Methods). Tables 1 and 2 present the mean, standard deviation (SD) and violin plots of the distribution of 1-ADRDI and Naive ProtoDrift (NP), ProtoDrift without optimised weights, in our two hospitals and in each of their subgroups. Although 1-ADRDI and NP share similar means across most subgroups, their distinct distributions suggest that each metric capture different aspect of treatment adjustments.

**Table 1:**
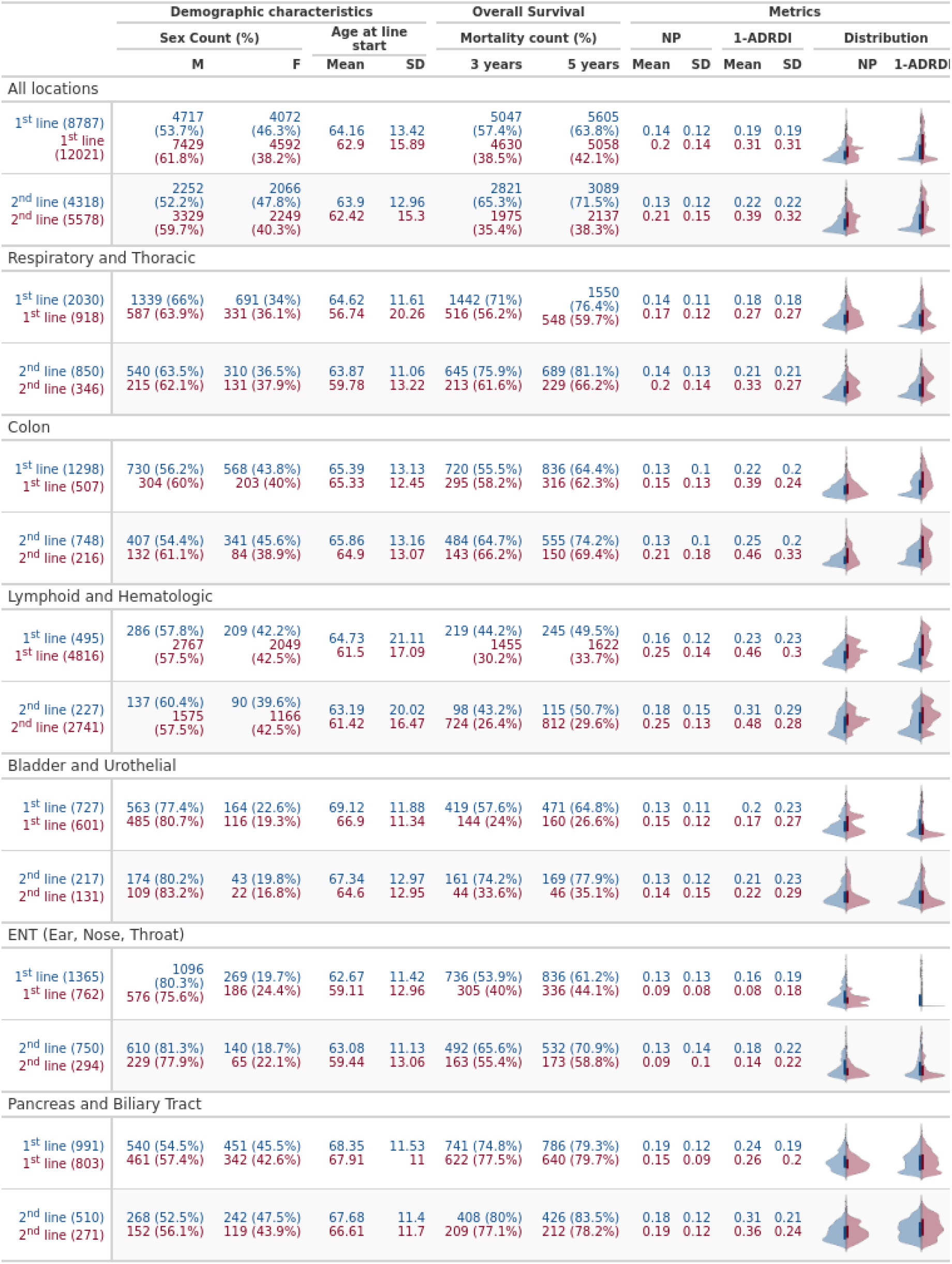
Demographic, overall survival, and distribution of ProtoDrift (NP) and 1-ADRDI in our two cohorts and each group of the study. 1-ADRDI is used to facilitate comparison with ProtoDrift (NP), as ADRDI is high when treatment closely aligns with the planned protocol, whereas ProtoDrift (NP) is low. Blue is used for data of the first hospital (HEGP) and red for the second (UHB) (1/2)

**Table 2:** Demographic, overall survival, and distribution of ProtoDrift (NP) and 1-ADRDI in our two cohorts and each group of the study. 1-ADRDI is used to facilitate comparison with ProtoDrift (NP), as ADRDI is high when treatment closely aligns with the planned protocol, whereas ProtoDrift (NP) is low. Blue is used for data of the first hospital (HEGP) and red for the second (UHB) (2/2)

|  | Demographic characteristics |  |  |  | Overall Survival |  | Metrics |  |  |  |  |  |
| --- | --- | --- | --- | --- | --- | --- | --- | --- | --- | --- | --- | --- |
|  | Sex Count (%) |  | Age at line start |  | Mortality count (%) |  | NP |  | 1-ADRFDI |  | Distribution |  |
|  | M | F | Mean | SD | 3 years | 5 years | Mean | SD | Mean | SD | NP | 1-ADRFDI |
| Breast |  |  |  |  |  |  |  |  |  |  |  |  |
| 1<sup>st</sup> line (1235) | 51 (4.1%) | 1184 (95.9%) | 57.27 | 13.71 | 398 (32.2%) | 486 (39.4%) | 0.09 | 0.09 | 0.11 | 0.17 |  |  |
| 2<sup>nd</sup> line (563) | 23 (4.1%) | 540 (95.9%) | 57.36 | 13.16 | 258 (45.8%) | 296 (52.6%) | 0.1 | 0.11 | 0.16 | 0.19 |  |  |
| Ovary |  |  |  |  |  |  |  |  |  |  |  |  |
| 1<sup>st</sup> line (502) | 0 (0%) | 502 (100%) | 65.42 | 13.13 | 251 (50%) | 291 (58%) | 0.17 | 0.14 | 0.16 | 0.15 |  |  |
| 2<sup>nd</sup> line (313) | 0 (0%) | 313 (100%) | 66.22 | 11.69 | 167 (53.4%) | 199 (63.6%) | 0.12 | 0.11 | 0.15 | 0.17 |  |  |
| Stomach |  |  |  |  |  |  |  |  |  |  |  |  |
| 1<sup>st</sup> line (433) | 279 (64.4%) | 154 (35.6%) | 62.73 | 13.5 | 288 (66.5%) | 302 (69.7%) | 0.13 | 0.09 | 0.23 | 0.2 |  |  |
| 2<sup>nd</sup> line (207) | 135 (65.2%) | 72 (34.8%) | 61.75 | 13.1 | 150 (72.5%) | 156 (75.4%) | 0.13 | 0.1 | 0.24 | 0.21 |  |  |
| Liver |  |  |  |  |  |  |  |  |  |  |  |  |
| 1<sup>st</sup> line (848) | 723 (85.3%) | 125 (14.7%) | 68.3 | 10.1 | 391 (46.1%) | 451 (53.2%) | 0.22 | 0.11 | 0.16 | 0.28 |  |  |
| 2<sup>nd</sup> line (152) | 131 (86.2%) | 21 (13.8%) | 70.47 | 9.68 | 74 (48.7%) | 80 (52.6%) | 0.21 | 0.17 | 0.36 | 0.36 |  |  |
| Neurology |  |  |  |  |  |  |  |  |  |  |  |  |
| 1<sup>st</sup> line (685) | 317 (46.3%) | 368 (53.7%) | 54.65 | 17.23 | 52 (7.6%) | 66 (9.6%) | 0.27 | 0.13 | 0.44 | 0.41 |  |  |
| 2<sup>nd</sup> line (411) | 198 (48.2%) | 213 (51.8%) | 54.8 | 16.01 | 27 (6.6%) | 28 (6.8%) | 0.27 | 0.17 | 0.54 | 0.41 |  |  |
| Melanoma |  |  |  |  |  |  |  |  |  |  |  |  |
| 1<sup>st</sup> line (1473) | 882 (59.9%) | 591 (40.1%) | 67.04 | 14.57 | 689 (46.8%) | 731 (49.6%) | 0.13 | 0.14 | 0.05 | 0.11 |  |  |
| 2<sup>nd</sup> line (628) | 371 (59.1%) | 257 (40.9%) | 65.84 | 14.07 | 274 (43.6%) | 295 (47%) | 0.1 | 0.14 | 0.07 | 0.14 |  |  |
| Myeloma |  |  |  |  |  |  |  |  |  |  |  |  |
| 1<sup>st</sup> line (609) | 328 (53.9%) | 281 (46.1%) | 67.22 | 10.84 | 162 (26.6%) | 189 (31%) | 0.18 | 0.13 | 0.35 | 0.27 |  |  |
| 2<sup>nd</sup> line (389) | 217 (55.8%) | 172 (44.2%) | 68.43 | 10.77 | 105 (27%) | 113 (29%) | 0.21 | 0.13 | 0.46 | 0.26 |  |  |

### 2.2 Optimised ProtoDrift Results

The optimisation of ProtoDrift weights with Cox regression (OP_Cox_) results in notable improvements of the C-index with regards to ADRDI across 16 out of 18 (89%) groups at HEGP. In 13 of them (72%) C-index increases with at least 1%. This trend is also observed when optimising the weights with the logistic regression (OP_Logit_), with gains in 15 groups (83%), and gains of at least 1% in 9 groups (50%). These results are summarised in Figure 2.

**Figure 2:**
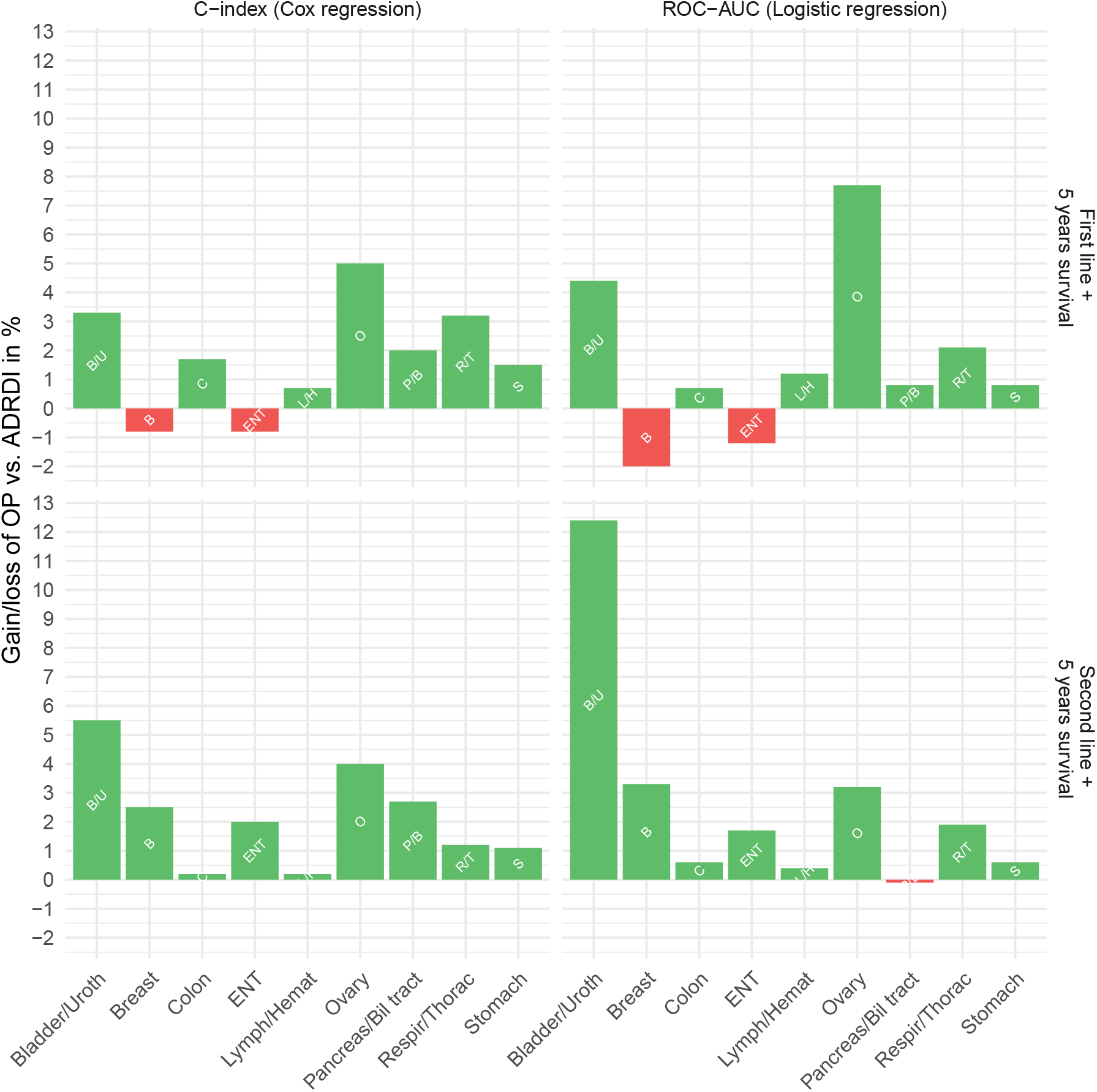
5-year overall survival prediction score gains when using OP_Cox_ (on the left) or OP_logit_ (on the right) *vs*. ADRDI. Relative gains are in green, losses are in red.

We observe a smaller SD of gains with OP_Cox_ with regards to OP_Logit_ that reaches the higher gains with over 7*·*5% and 12% for Ovary, first line and Bladder/Urothelial, second line, respectively.

### 2.3 Naïve ProtoDrift results

To evaluate the performance of Naïve ProtoDrift (NP) against the baseline ADRDI, we computed and compared three Cox-based performance indicators: significance of the regression coefficient, C-index, and LogRank p-values (see section 4.3 in Methods).

We find that NP performs comparably or better than ADRDI in a substantial proportion of patient groups. Specifically, 50% of groups at both HEGP and UHB (9/18 and 10/20, respectively) show at least one performance gain and no loss across the three indicators. Overall, 67% of HEGP groups and 75% of UHB groups have either comparable or improved results with NP.

Notably, a subset of groups demonstrate consistent improvements across all three indicators. At HEGP, these include Respiratory/Thoracic, Colon, and Ovary (first line), as well as ENT (second line). At UHB, they include Colon, ENT, and Melanoma (first line), and Colon and Melanoma (second line). Conversely, ADRDI outperforms NP (i.e., at least one loss and no gain) in 18% of the total groups.

These results suggest that ProtoDrift, even without optimisation, offers a more nuanced capture of treatment adherence compared to ADRDI.

For a visual overview, we refer the reader to the UpSet-style summaries and per-group bar plots provided in Appendix Figures 10 and 11, which synthesize the comparative performance across patient subgroups. For full transparency, the complete numerical results—covering all three Cox-based indicators and additional logistic regression metrics—are available in Appendix Tables B.3

### 2.4 Illustration with the prediction of the 5-year overall survival in the Respiratory/Thoracic group

NP and OP have been computed for each group of the first hospital (HEGP), and in one group only (Respiratory/Thoracic) in the second hospital for external validation. The complete results of the comparative analysis are available at https://files.inria.fr/protodrift-surv/. Here, we focus on results obtained on the illustrative group for which ProtoDrift was optimised in both hospitals: patients with Respiratory/Thoracic cancer, in their first line of treatment, using Cox regression prediction.

Figure 3 shows that, in patients treated in HEGP hospital, both NP and OP_Cox_ models demonstrate superior performance to ADRDI across the three considered indicators (see Figure 3b). Firstly, both NP and OP_Cox_ show significant contributions to the Cox regression models (p-values*<*0.01 and =0.01, respectively), indicating a reliable prediction of the mortality risk, which is not observed with ADRDI (p-value=0.23). Secondly, the two models outperform ADRDI in predictive performance, evidenced by higher C-index scores. Lastly, the discriminative power analysis, using Kaplan-Meier survival curves (see Figure 3a), confirmed that OP_Cox_, and to a lower extend NP, effectively differentiate between patient survival outcomes across quartiles, validated by significant LogRank test results (see Figure 3b). Interestingly, we observe with OP_Cox_, which best align with the outcomes, an inversion of second and first quartiles that can not be observed with other approaches.

**Figure 3:**
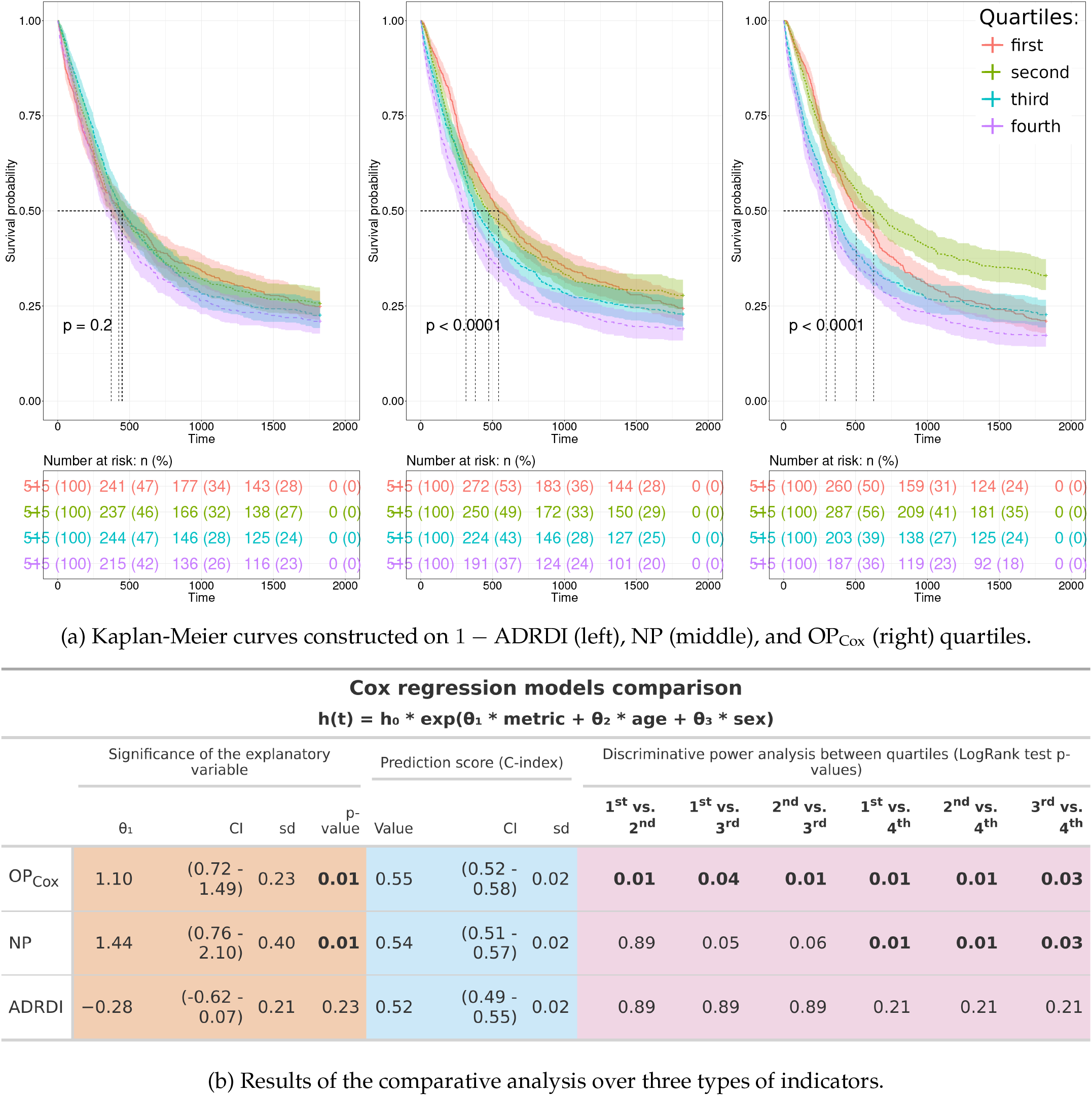
Comparative performances of the prediction with the Cox regression model of the 5-year overall survival at the end of the first line of the Respiratory/Thoracic group at the HEGP hospital.

Figure 4 shows that in patients treated in UHB, none of the metric shows a significant contribution in the Cox regressions models (see Figure 4b). However, we qualitatively observe that the Kaplan Meier curves are more spaced apart with NP or OP_Cox_ than with ADRDI quartiles, underscoring a better overall discriminative power (see Figure 4a).

**Figure 4:**
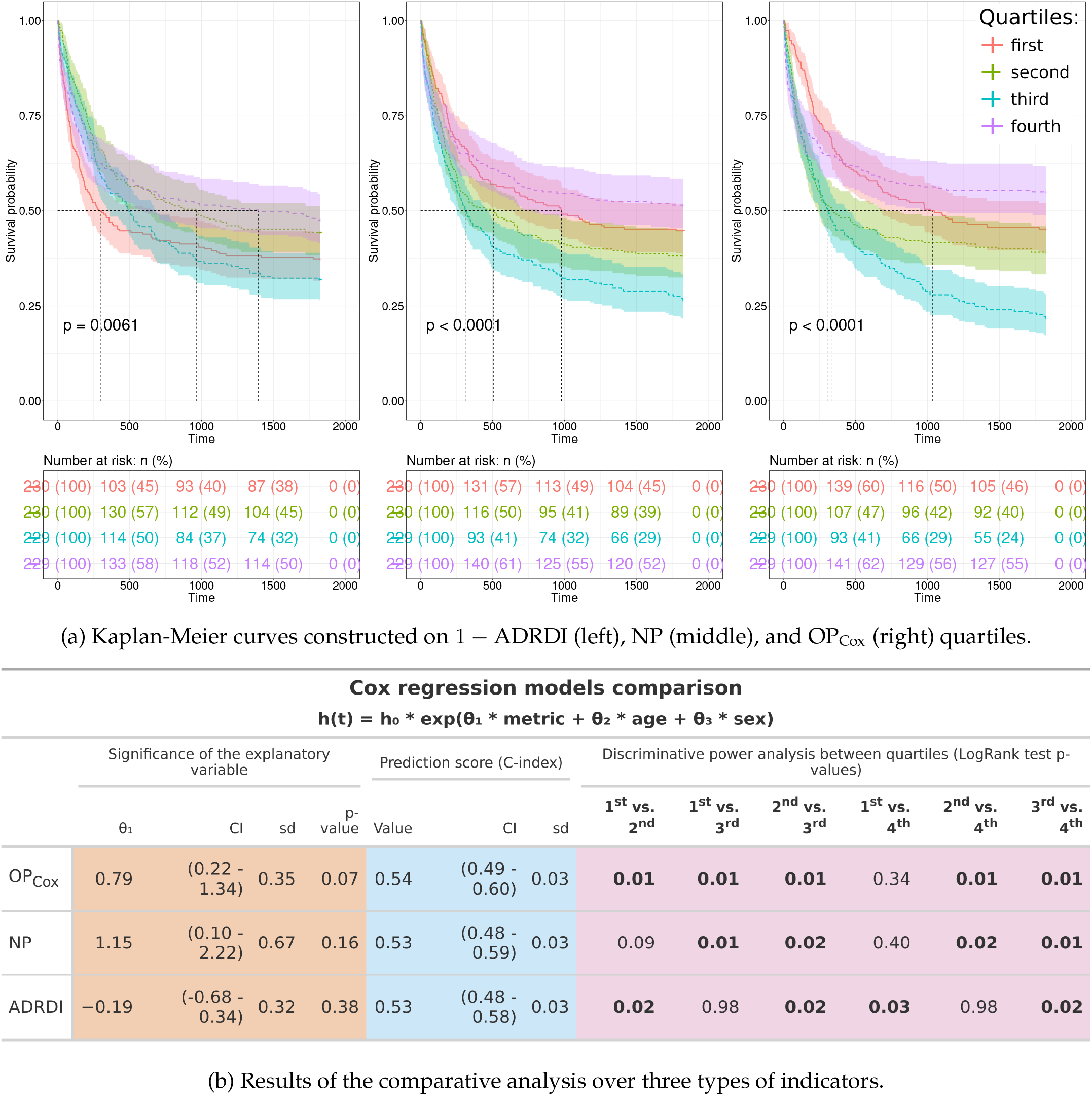
Comparative performances of the prediction with the Cox regression model of the overall survival at the end of the first line of the Respiratory/Thoracic group at the UHB hospital.

### 2.5 Relative importance of ProtoDrift weights

We computed heat maps to provide for each group a view of the relative importance of the weights of ProtoDrift, and thus of the various types of adjustment (*e*.*g*., dose, timing, cycle delay). Figure 5 shows that for the Respiratory/Thoracic, first line group, both at HEGP and UHB, best survival prediction is reach for high value of *α* and low value of *β, i*.*e*., when *ω*_*t*_ *> ω*_*d*_ and *ω*_*intra*_ *> ω*_*inter*_. In other terms, in-cycle time changes have more impact than in-cycle dose changes and changes in in-cycle timing and dosing have more impact than cycle delays (see Figure 7). We observe a similar trend for other groups, such as ENT, second line or Pancreas/Biliary Tract, first line (see https://files.inria.fr/protodrift-surv/year-survival-22 and https://files.inria.fr/protodrift-surv/year-survival-32). However, in other groups, other balances are observed. For example, in Ovary, second line, and Colon, first line, we observe a stronger impact of dose changes with regards to time changes (see https://files.inria.fr/protodrift-surv/year-survival-30 and https://files.inria.fr/protodrift-surv/year-survival-16).

**Figure 5:**
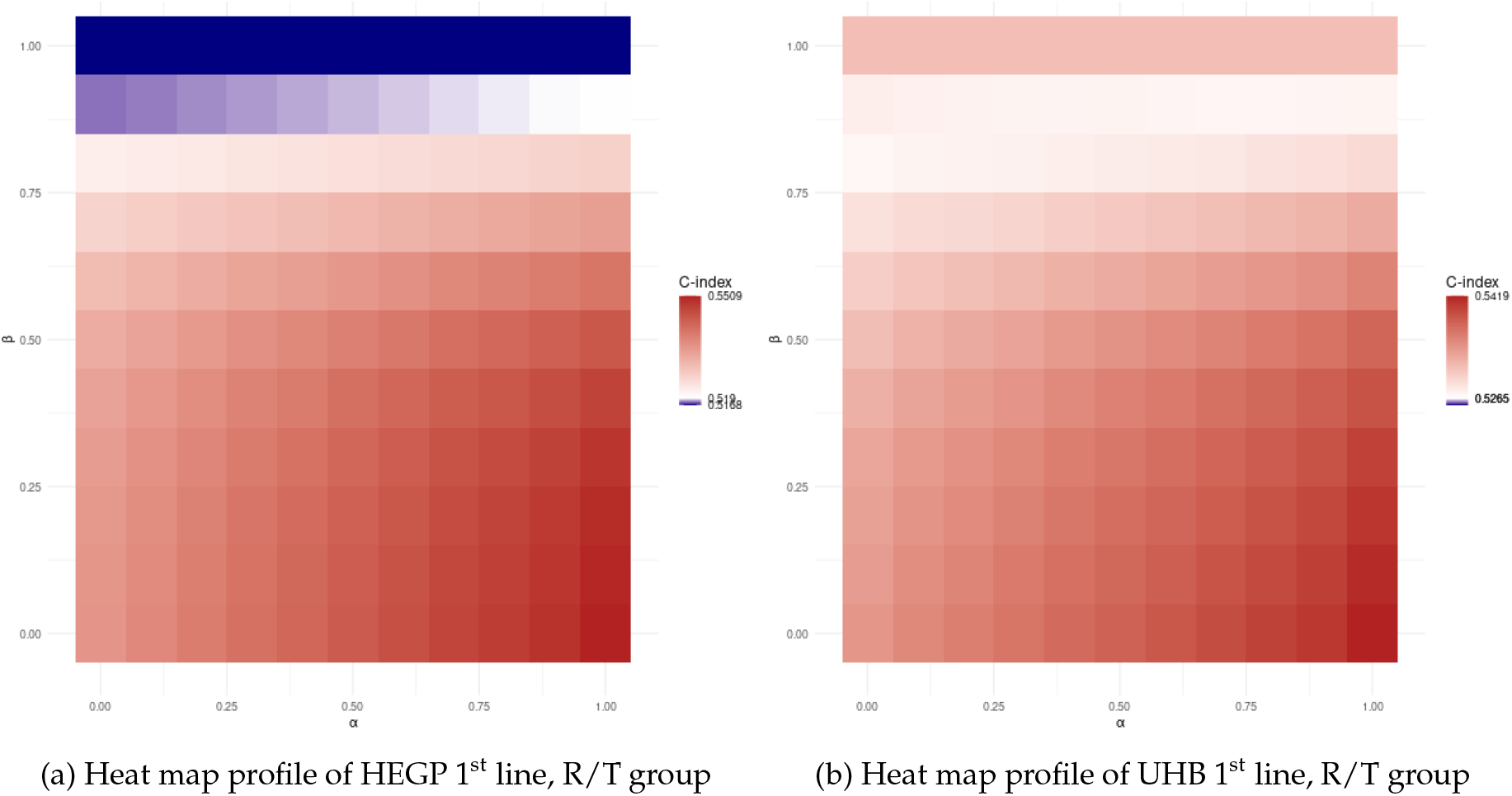
Heat map representation of prediction gains with ProtoDrift (OP_Cox_) *vs*. ADRDI depending on values of *α* (i.e,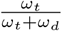) and *β* (i.e, 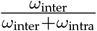), in patients with Respiratory/Thoracic (R/T) cancer groups in their first line of treatment, either at HEGP (left) or UHB (right). Figure 7 provides an interpretation guide to these heat maps.

## 3 Discussion

We introduced ProtoDrift, a novel metric for the fine-grained measure of chemotherapy adherence, and demonstrated its superiority to the state-of-the-art RDI, by showing how Protodrift, in particular when optimised, is more significantly associated with clinical outcomes.

This establishes our metric a new foundational tool to customise the design of chemotherapy regimens and the clinical adjustments of chemotherapy.

The comparative analysis of ProtoDrift weights reveals that ProtoDrift particularly outperforms ADRDI when the timing weight (*ω*_*t*_) exceeds the dosage one (*ω*_*d*_). This can be explained by the fact that temporal adjustments are loosely considered in ADRDI that sums the dose changes for a defined period of time, thus do not consider punctual timing adjustments.

Moreover, the relative importance of ProtoDrift weight provides elements of interpretability about the type of adjustment that might impact clinical outcomes. For instance, Figure 5 illustrates that in the first line treatment of Respiratory/Thoracic cancer, an overall delay in cycles or a change in dosage may have minimal impact on survival compared to an in-cycle administration time shift. Such distinctions could not be observed when using ADRDI.

ProtoDrift also brings new insights by its ability to better discriminate between patient groups. Kaplan-Meier analyses, Figures 3a and 4a) enable to discuss the hypothesis that better adherence to treatment correlates with improved survival. Indeed, at UHB, the least adherent quartile shows to be significantly associated with higher survival rates. This observation echoes studies about chemotherapy management in elderly patients, particularly those with non-small cell lung cancer (NSCLC), where reduced dosages and delayed administrations might benefit survival.^18^ This highlights the potential of ProtoDrift to further uncover nuanced treatment impacts, with high potential in advancing precision medicine in cancer therapy.

We analysed this discriminative power using quartiles based on OP, NP and 1-ADRDI. In contrast, many RDI studies split their cohorts into two groups using an 80% threshold, a less stringent division. Our quartile division, although generally robust, is not always suitable for discerning patient clusters. This limitation is observed in Figure 11 where certain cohorts gain in C-index but looses in discriminative power such as HEGP Bladder/Urothelial in first line (see https://files.inria.fr/protodrift-surv/year-survival-8).

It is worth noting that ProtoDrift can find applications beyond the field of oncology, and particularly suits other treatments associated with cyclic protocols.^19^ On a group of patients with Auto-immune and Inflammatory diseases, not included in our study, the comparison of ProtoDrift to RDI shows gains on our three performance indicators (see https://files.inria.fr/protodrift-surv/year-survival-4).

ProtoDrift can be explored further. In particular, its interpretability capabilities could be extended by considering the relative weights of the molecules of a regimen (*i*.*e*., does a dose change with molecule A will greatly impact the outcome than a dose change of molecule B?), or the mode of administration (*i*.*e*., does a bolus administration will greatly impact than slow infusion of the same molecule?). However, one can expect that adding more parameters will complexify interpretations. In assessing ProtoDrift, we avoided combining patient groups from the two hospitals or directly applying optimised weights from one hospital to the other on purpose. This strategy was selected to respect the unique characteristics and treatment deviations inherent to each hospital population. Our approach focuses on understanding specific hospital contexts rather than creating a universally applicable model. This distinction supports the use of ProtoDrift to explore variability in treatment adherence and its impact, tailored to each clinical setting. This highlights that ProtoDrift does not aim at being a predictive tool, but a means to assess and visualise the impact of different types of adjustment on clinical outcomes.

The reproducibility of our observations with ProtoDrift could be further extended as they were limited to seven groups for NP (Figure 11) and one group for OP (Figures 3 and 4). In this work, we optimised ProtoDrift for only one UHB location, primarily due to constraints on computational time and data access. Particularly, the weight optimisation, which involves fitting and predicting regression models multiple times, is time-consuming. We aimed at additional validation and interoperability with our effort to provide an open implementation of ProtoDrift that takes as input treatment descriptions in an easy-to-reuse tabular format (see Code availability section for details).

## 4 Methods

### 4.1 Data sources and study design

We independently collected chemotherapy prescriptions and administrations HEGP and UHB hospitals from 2003-07-01 to 2021-12-15 (about 18.5 years).^20^

Both hospitals include in-stay and after-stay patient survival data that is the outcome used in this study.^21^

We relied on ChemoOnto for reconstructing chemotherapy treatment course from anti-cancer medication records and theoretical regimens.^22,23^

Groups of patients were defined by tumour location. In each hospital, only groups with at least 400 patients were considered. For each location, we also distinguished between chemotherapy administrated in either the first or second line of treatment.

As illustrated by table 6, we excluded 2,944 patients at HEGP and 11,428 at UHB because of poor data quality or inconsistency. 13 locations with more then 400 patients were kept, six found both at HEGP and UHB, three at HEGP only, and four at UHB only. When considering treatment lines, the study is composed of 18 groups from HEGP and 20 groups from UHB, described in tables 1 and 2. Tumour location groups shown in Figure 6 are not disjoint because patients may have more than one cancer location. For each location group, all patients have followed a first line of treatment, and potentially, but not always, a second. Consequently, the size of the second line groups is smaller than that of the first line groups, as can be observed in tables 1 and 2.

**Figure 6:**
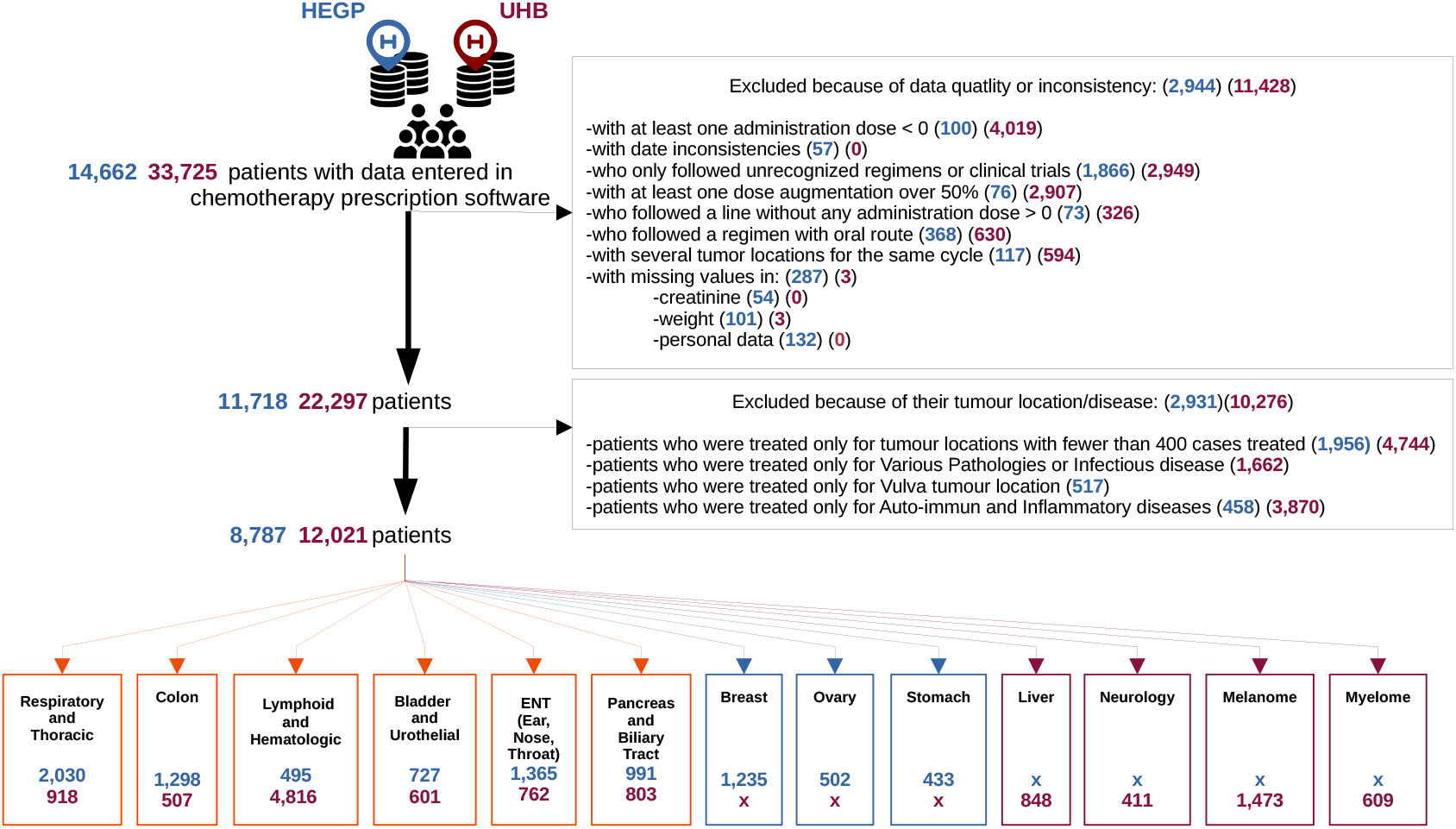
Study profile. After data cleaning, we considered tumour locations with more than 400 cases treated (n*>*400). Six cancer locations are common to our two hospitals, three locations from HEGP only, and four from UHB only.

**Figure 7:**
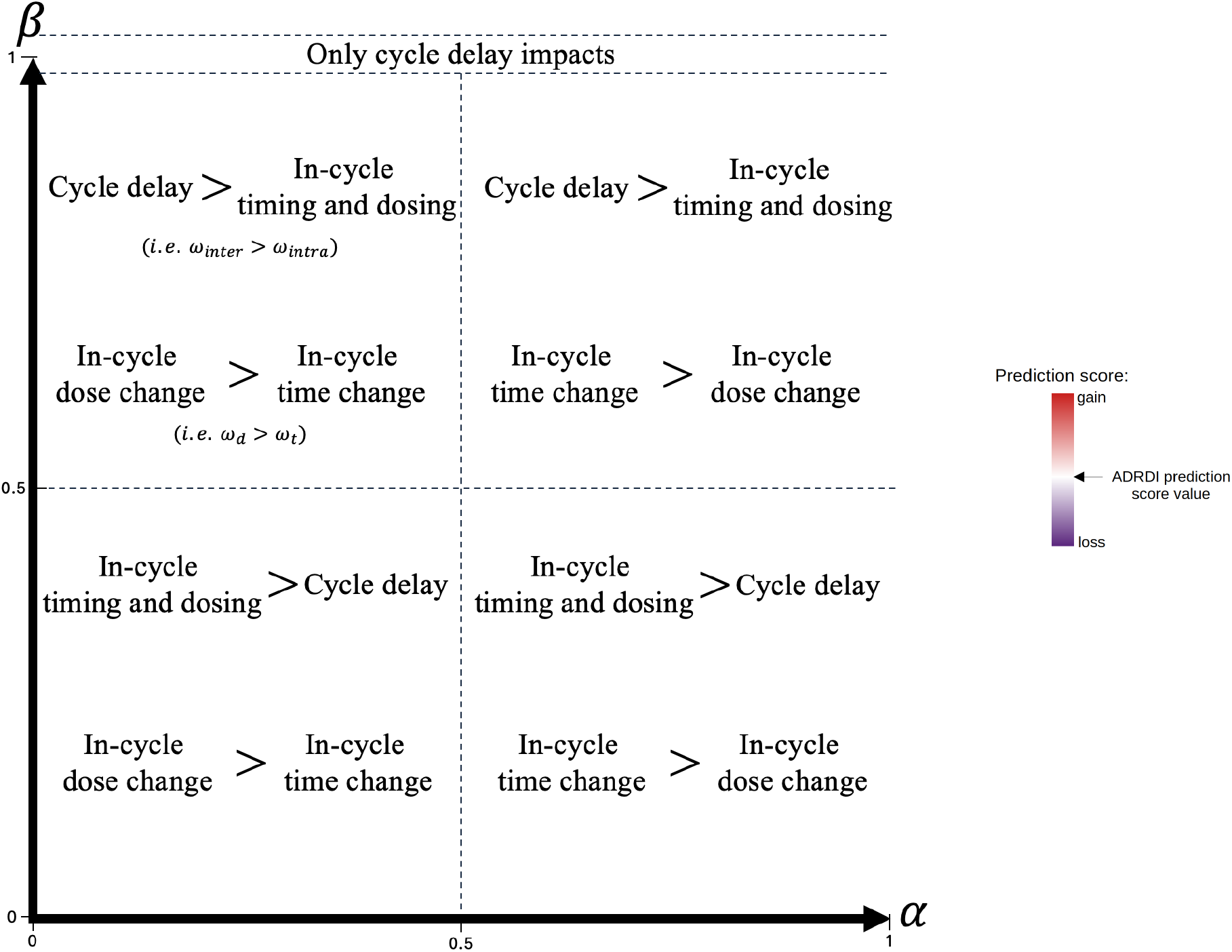
Interpretation guide for the relative importance of various ProtoDrift parameters on survival prediction.

Details about patients selection are provided in Appendix B.1.

### 4.2 ProtoDrift model development

#### 4.2.1 Definition of ProtoDrift

ProtoDrift is a multi-level weighted mean designed to quantify the deviation between an administered chemotherapy treatment and the initially prescribed treatment protocol. Possible deviations are quantified with dissimilarities, scaled from 0 to 1. Values close to 1 indicate high deviation.

The first step of ProtoDrift is to compute dissimilarities at the administration level, between actual and theoretical cycle. For each administration, a dissimilarity *δ*_adm_ is computed as a weighted mean of differences in dose (*δ*_*d*_) and timing (*δ*_*t*_) from the theoretical cycle. Formally,

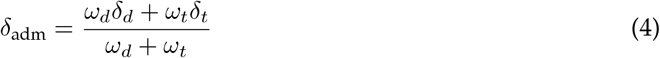

where *ω*_*d*_ and *ω*_*t*_ are the weights associated with dose and time changes. An alignment algorithm pairs the planned and actual administrations to ensure that ProtoDrift consider them only once. Details of the alignment algorithm are documented in Appendix Figure 8 and pseudo-code 1.

**Figure 8:**
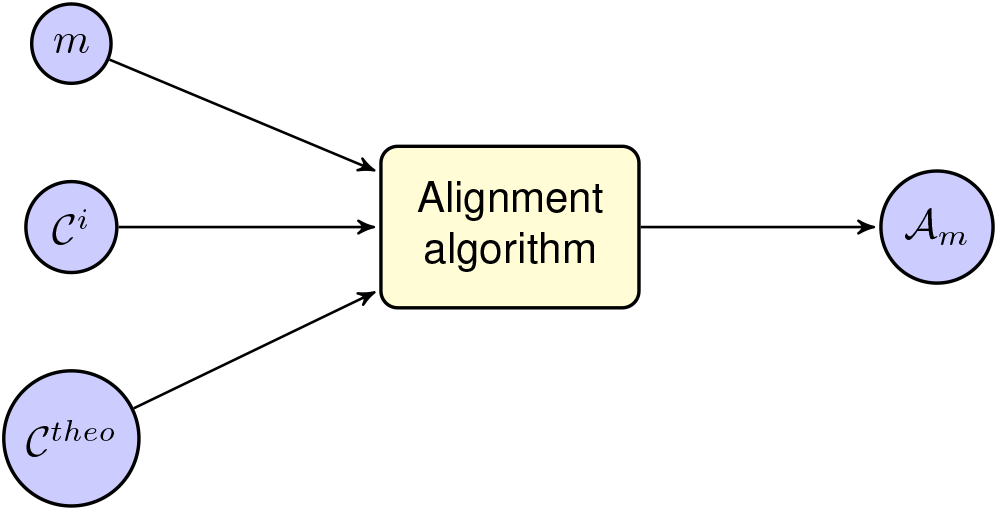
Schematic view of the input and output of the ProtoDrift alignment algorithm, with *𝒞*^*theo*^ and *𝒞*^*i*^ a theoretical and an actually administrated cycle, *m* a pair (molecule, mode of administration) and *𝒜*_*m*_ the set of aligned pairs of administrations for *m*.

Administration-level dissimilarities are then averaged at the molecule level (*δ*_*m*_), by considering all the administrations of a same drug within a cycle. Next, an intra-cycle dissimilarity, noted *δ*_intra_, is calculated by aggregating molecule-level dissimilarities within a cycle. Besides, ProtoDrift computes an inter-cycle dissimilarity, noted *δ*_inter_ that measures delays (or rare advances) in starting new cycles. Then, intra- and inter-cycle dissimilarities are aggregated at the cycle level with a weighted mean as follow:

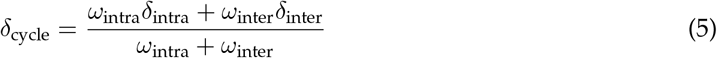

where *ω*_intra_ and *ω*_inter_ are the respective weights for the intra- and inter-cycle dissimilarities. Finally, cycle-level dissimilarities are summed at the line level to provide the final metric named ProtoDrift:

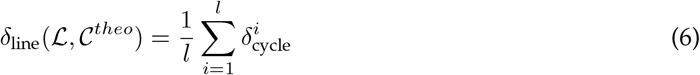

where *l* is the number of cycles of the treatment line *ℒ*, and *𝒞* ^*theo*^ the theoretical cycle as defined in the protocol. Appendix Figure 9 summarises the various levels of dissimilarity and aggregation. We measure ProtoDrift at the level of the treatment line (*δ*_line_). Firstly because we want to account for adjustments occurring within a treatment line. Secondly because studies with RDI generally compute its value for a treatment line. Details on the computation of ProtoDrift are provided in Appendix A.1.2.

**Figure 9:**
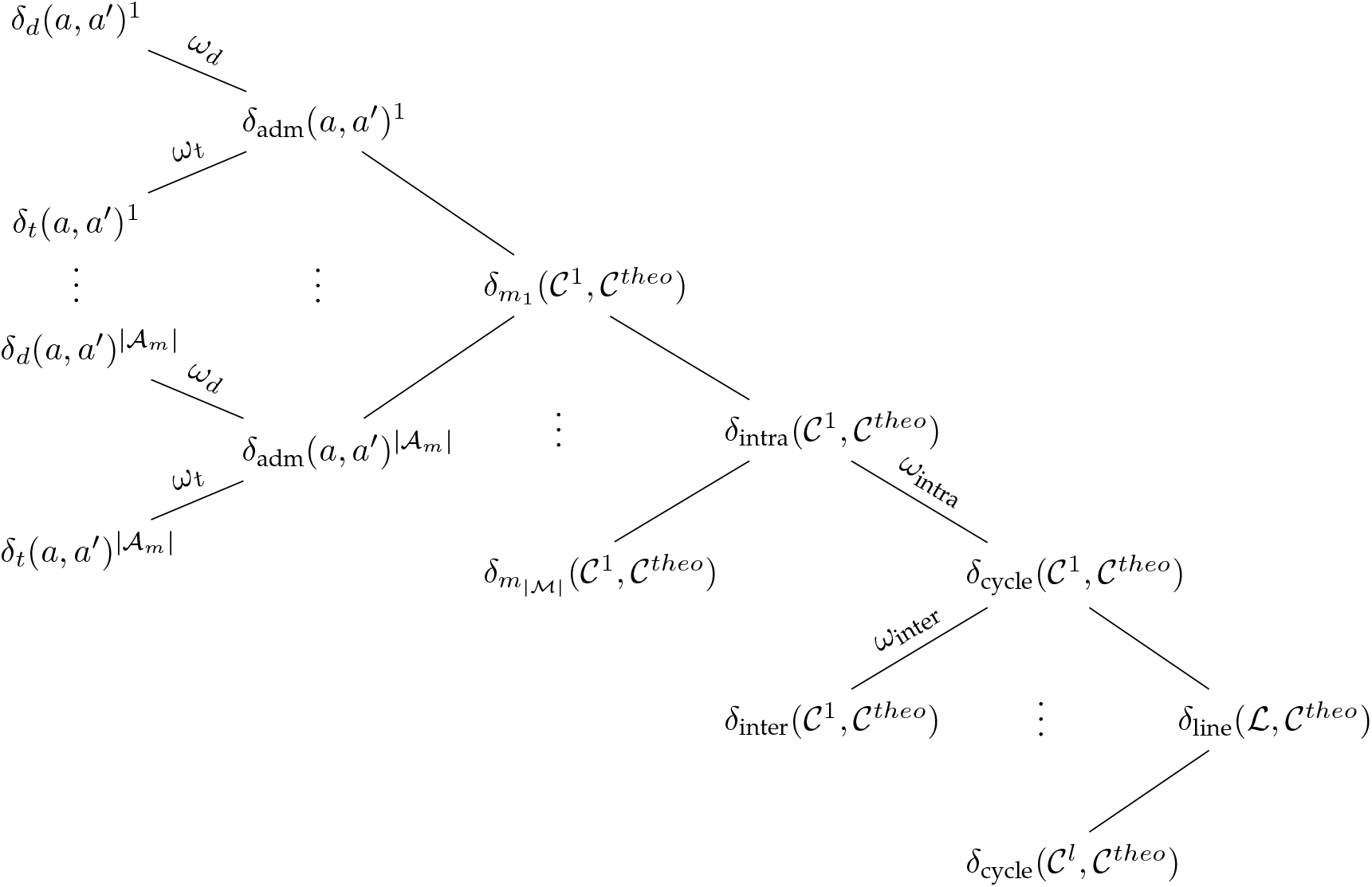
The weighted dissimilarities of ProtoDrift, at different levels, from dose and timing dissimilarities of single administrations, at the extreme left, passing by molecule level dissimlarities, intra- and inter-cycle dissimilarities, to finally aggregate at the cycle and line levels, at the extreme right. Dissimilarities measures the gap between a theoretical protocol defined by a theoretical cycle *𝒞*^*theo*^ and an actual line of treatment *ℒ*composed of *l* cycles followed by a patient. (*a, a*^*′*^) are pairs of administrations, one actual and one planned, compared to compute dose and time shifts. These pairs are computed by the alignment algorithm described figure 8. |*𝒜*_*m*_| is the maximum number of administrations of either the actual or theoretical cycle for the molecule *m*. |*M*| is to the number of distinct anti-cancer molecules in the cycles.

**Figure 10:**
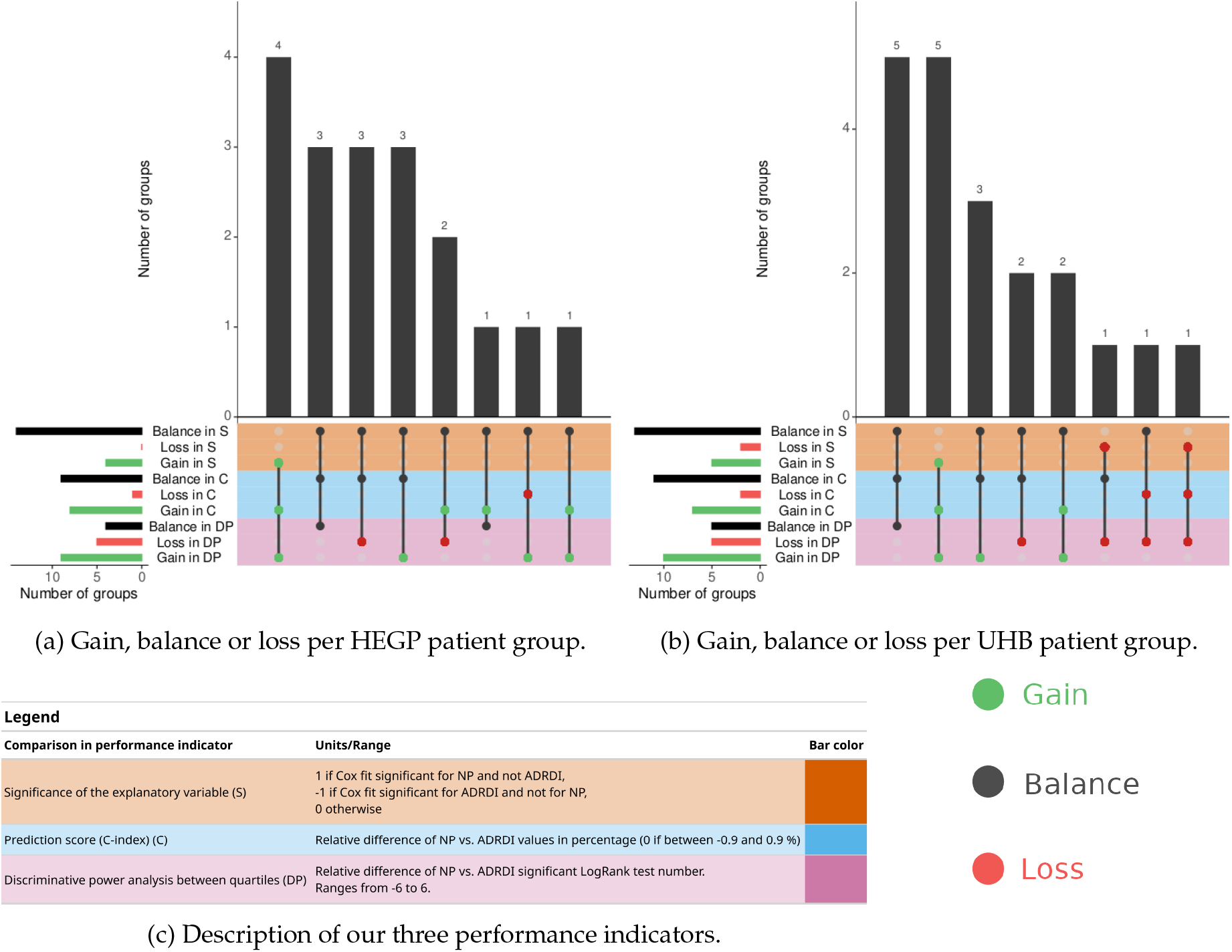
Overall advantage of using Naïve ProtoDrift (NP) *vs*. ADRDI, across the 18 patient groups of HEGP (a) and the 20 of UHB (b). Vertical bars quantify the number of groups that either gain, loss or have balanced results in using NP instead of ADRDI with regards to three performance indicators, described in the colour-coded matrix (c). Green indicate gains, black indicates balanced indicators, and red losses. We present here performances obtained on 5-year overall survival.

**Figure 11:**
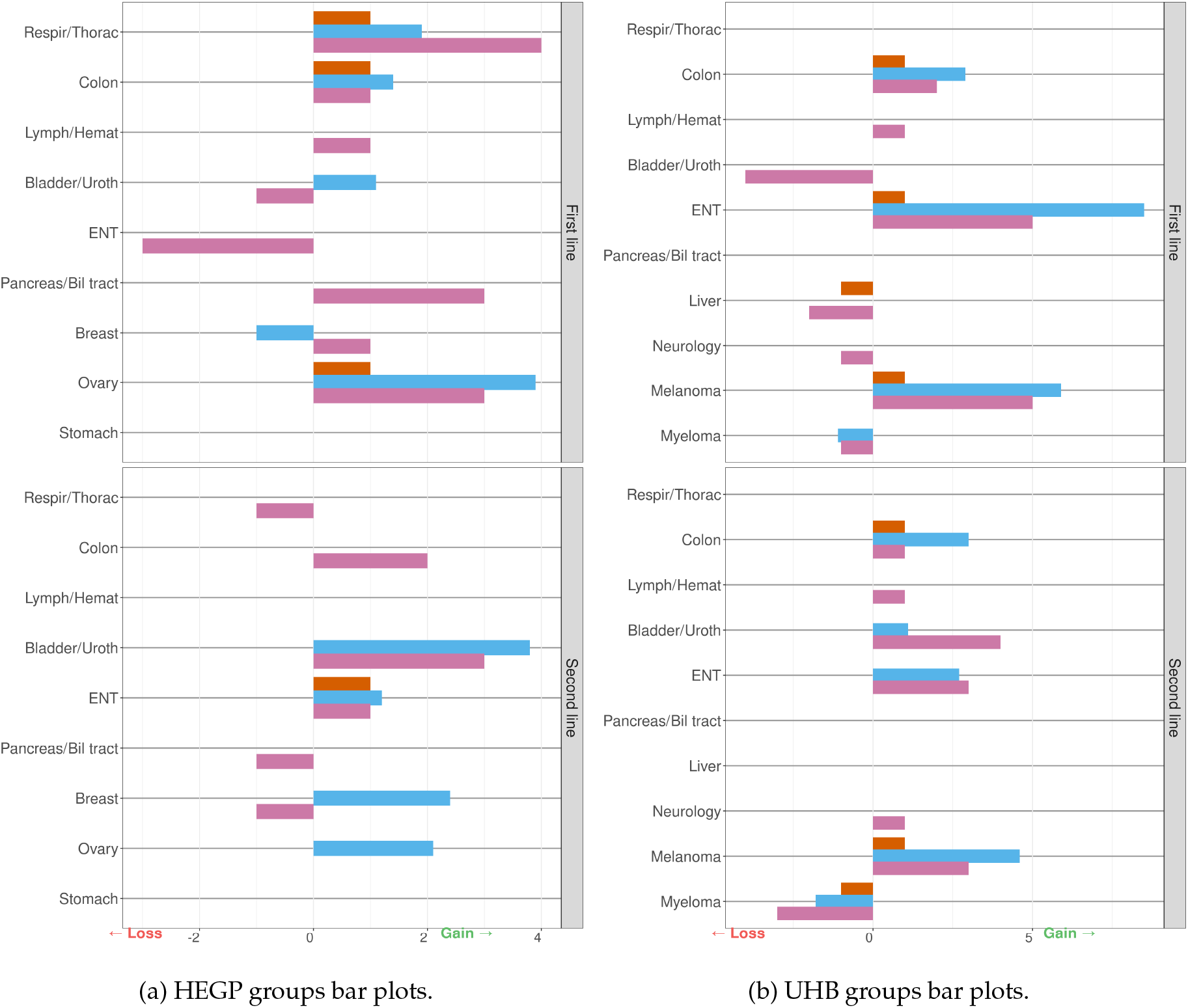
Detailed advantages of using NP in comparison with ADRDI, per cancer location and first or second line of treatment across the 38 groups of HEGP (a) and UHB (b). Each color bar refers to one of the three indicators presented Figure 10 (c) for a group. Bars to the right indicate gains and to the left indicate losses for the three indicators. Note that each indicator has a different unit. No bar indicates a balanced indicator for the group.

#### 4.2.2 Optimisation of ProtoDrift weights

ProtoDrift associates a weight with each of its composing dissimilarities, each one reflecting the relative importance of a type of deviation to a chemotherapy protocol. We particularly focused on the weights that reflect dose adjustments, drug removal and timing shifts, either within or between cycles (delays), *i*.*e. ω*_*d*_, *ω*_*t*_, *ω*_intra_ and *ω*_inter_, associated with *δ*_*d*_, *δ*_*t*_, *δ*_intra_ and *δ*_inter_ dissimilarities, respectively.

We distinguish two versions of ProtoDrift: *Naive ProtoDrift (NP)* where each weight is set to 1, *i*.*e*., each type of adjustment has a similar weight; and *Optimised ProtoDrift (OP)* where optimal weights are set on the basis of a grid search. This optimisation targets two goals:

- identifying the weight combination that offers the best survival predictions,
- helping interpreting what type of adjustment (*e*.*g*., dose or time adjustment) has more impact on survival outcomes.

Optimal weight combinations can be searched for various groups, such as different cancer locations. The optimisation is performed on the 3- and 5-year overall survival, but it could alternatively be on progression free survival or toxicity occurrences. Here, we preferred overall survival as it is recorded routinely and with good quality. Moreover, RDI studies we compare with, usually takes the overall survival as their main outcome.

To simplify the exploration for optimal weights, we define two quantities *α* and *β*, which values are between 0 and 1, that summarise the four weights considered for optimisation (*ω*_*d*_, *ω*_*t*_, *ω*_intra_, *ω*_inter_):

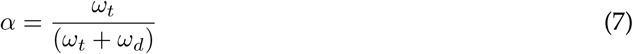

and

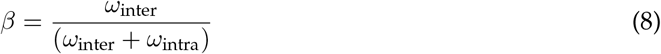

Those enable to rewrite the two following components of ProtoDrift:

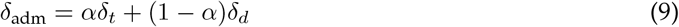

and

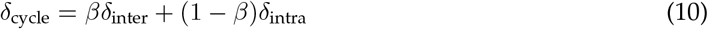

(See Appendix A.2 for the proof).

We grid search over *α* and *β* by a step of 0.1 computing at each step ProtoDrift (*δ*_line_) and fit a logistic regression and a Cox regression model according to the two following equations:

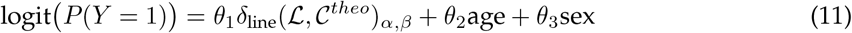

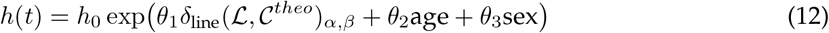

In the logistic regression (11), the binary response variable Y encodes the occurrence of the event before the survival time. In Cox regression (12), *h* is the hazard function. For genital cancer locations, the last term is removed from both formulas (*θ*_3_sex).

Predictions are evaluated using ROC-AUC for logistic regression and C-index for Cox regression to select the best *α* and *β* values. We note OP_logit_ and OP_Cox_ the versions of Protodrift optimised either with the logistic or the Cox regression. We use bootstrap resampling with 500 samples to account for the variability in the sampled populations and assess the uncertainty of predictions.

The grid search computes a prediction score for every combinations of (*α, β*) pairs, what provides a view of their relative importance for the prediction of survival, and can be interpreted as:

- a high value of *α* (close to 1) signifies that *ω*_*t*_ *> ω*_*d*_, that is to say in-cycle timing of administrations has more impact than dose changes;
- a high value of *β* (close to 1) signifies that *ω*_inter_ *> ω*_intra_, that is to say the overall cycle delay has more impact than the in-cycle timing and dose changes.

For a particular group of patients, this relative importance can be visualised with heat maps of the survival prediction score plotted according to *α* and *β* values. Results can be interpreted following the guidelines provided in Figure 7. When *β* = 1, ProtoDrift is constant, and so is the prediction score. Indeed, in this case, only the overall cycle delay impacts (see proof in Appendix A.2).

### 4.3 Comparative analysis of ProtoDrift and Relative Dose Intensity

With the objective of demonstrating the benefit of ProtoDrift over the all drugs RDI (ADRDI), we compare their performance in predicting the overall survival with both logistic and Cox regression models. To this aim, we use either NP, OP or ADRDI as the explanatory variable of our two predictive models (11) and (12). The resulting models are detailed in Appendix A.2.

We assess the performance of the various approaches using three distinct indicators:

- The significance of the explanatory variable contribution to the regression model with the Wald test p-value associated with each approach.
- Prediction score comparison: Prediction performances are measured with the ROC-AUC for logistic regressions or C-index for Cox regressions. The difference of these metrics between models with ADRDI and NP, or between ADRDI and OP, gives the gain in performance of ADRDI *vs*. ProtoDrift.
- Discriminative power analysis: This evaluates the ability of each model to differentiate between four groups with distinct survival outcomes. First, we split patients into quartiles based on their values of NP, OP and 1-ADRDI. Second, by analysing Kaplan-Meier survival curves for these quartiles, we assess the effectiveness of the models in distinguishing varying survival outcomes, validated through LogRank tests for statistical significance. We work with 1-ADRDI, to facilitate the comparison with Protodrift, as ADRDI is high when a treatment is similar to the planned protocol, while Protodrift is low.

To assess the reproducibility and validity of ProtoDrift, we conducted comparative analyses using datasets from both HEGP and UHB. For the HEGP dataset, we performed comparisons between ADRDI *vs*. NP and ADRDI *vs*. OP across all patient groups. In contrast, for the UHB dataset, while the comparison between ADRDI *vs*. NP was conducted across all patient groups, the ADRDI *vs*. OP comparison was carried out for the Respiratory/Thoracic first line group only.

## Data Availability

All data produced in the present study are available upon reasonable request to the authors

https://files.inria.fr/protodrift-surv/

## 5 Data availability

ADRDI-NP-OP comparative analysis of the 18 HEGP patient groups, with both Cox and logistic regressions, and predicting both 3- and 5-year overall survival (72 analysis), are available at https://files.inria.fr/protodrift-surv/. Patient data were collected during healthcare, either at the Georges Pompidou Hospital in Paris, or at the University Hospital of Bordeaux, France. Data were used under the IRB CSE-21-16 and CER-BDX-2024–80, respectively. These personal data are not shared to preserve their confidentiality.

## 6 Code availability

The code to compute Naive ProtoDrift is available at https://gitlab.inria.fr/arogier/ `protodrift`. The code to optimise its weights is available at https://gitlab.inria.fr/arogier/protodrif The schema and the dictionary of the analysed data are those of ChemoOnto, available at https://gitlab.inria.fr/arogier/ChemoOntoTox.

## Acknowledgements

This work benefited from a grant from the joint Inria-Inserm PhD program, from Inria PhD support, and from a government grant managed by the Agence Nationale de la Recherche under the France 2030 program, reference ANR-22-PESN-0007, ShareFAIR. We extend our thanks to Juliette Murris, Mathilde Pezot, Victor Gondret and Jean Feydy for fruitful discussions.

## Declaration of generative AI in the writing process

During the preparation of this work, the authors used ChatGPT to improve language and readability. After using this service, the authors reviewed and edited the content as needed and take full responsibility for the content of the publication.

## 7 Author contributions

**AR:** Conceptualisation, Data curation, Formal analysis, Investigation, Methodology, Software, Visualisation, Writing - original draft, Writing - Review & Editing. **EA, EP:** Interpretation, Investigation, Discussion, Review & Editing. **CB, RG, VJ:** Data curation, Validation, Review & Editing. **BS, EZ:** Data curation, Interpretation, Investigation, Review & Editing. **BR:** Conceptualisation, Methodology, Validation, Resources, Writing - Review & Editing, Supervision, Project Administration, Funding acquisition. **AC:** Conceptualisation, Methodology, Validation, Resources, Writing - Review & Editing, Supervision, Project Administration, Funding acquisition.

## Appendix

### A Methods

#### A.1 ProtoDrift detailed methodology

##### A.1.1 Chemotherapy treatment concepts formalisation

- An *i*^th^ chemotherapy cycle followed is defined by a set of administrations *𝒞*^*i*^ = {*a*^1^, …, *a*^*n*^}. Each cycle has a duration 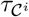. We denote by *𝒞*^*theo*^ the theoretical cycle defined by the regimen of duration 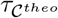.
- A line *ℒ* is an ordered set of followed cycles: *ℒ* = {*𝒞*^1^, …, *𝒞*^*l*^}. A line has a duration *τ* _*ℒ*_.
- A protocol is a repetition of theoretical cycles.
- An anti-cancer drug *m* is a pair *(molecule, mode of administration)*. For example, (cetuximab, continuous infusion) or (cetuximab, bolus).
- A drug administration *a* is composed of three dimensions: *⟨a*_*m*_, *a*_*t*_, *a*_*d*_*⟩*:
  – *a*_*m*_ is the anti-cancer drug *m* associated with the administration,
  – *a*_*t*_ is the time between the start of the cycle and the administration. For example, 7 if the administration must be done 7 days after the start of the cycle,
  – *a*_*d*_ is the dose. For example, 300 if the administration must be done with a dose of 300 mg.
- 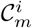 is the subset of administrations in the cycle *C*^*i*^ that have the same anti-cancer drug *m* (i.e., the same pair *(molecule, mode of administration)*).
- *ℳ* is the set of anti-cancer drugs in *𝒞*^*i*^ or *𝒞*^*theo*^:

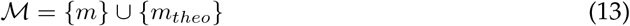

where {*m*} (respectively {*m*_*theo*_}) is the set of drugs in cycle *𝒞*^*i*^ (resp. *𝒞*^*theo*^).

###### ADRDI notation

Here we adapt ADRDI definition (formula 3), with concept formalisation of section A.1.1.

For an anti-cancer drug *m*, a line *ℒ*, its duration in weeks *τ*_*ℒ*_, we have:

- The dose-instensity DI (1) is defined as:

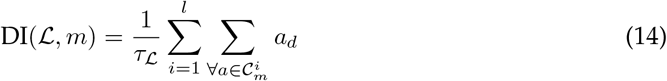
- The theoretical dose-intensity:

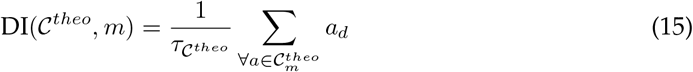
- The relative dose-intensity of line *ℒ*:

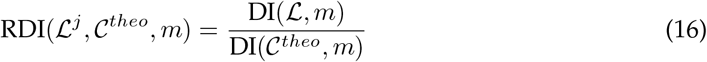

ADRDI across all drugs in a treatment line *ℒ* is defined as:

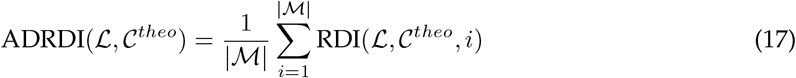

This ADRDI metric provides a normalised measure to compare the adherence of actual treatment to the prescribed regimen, with values ranging between 0 and 1. A higher ADRDI value indicates closer adherence to the prescribed treatment regimen, contrasting with the line-protocol dissimilarity: *δ*_*line*_ where a higher value signifies greater deviation.

##### A.1.2 The different dissimilarity components of ProtoDrift and their formulation

At different time scales of chemotherapy treatment, we define dissimilarities traducing deviations with the planned protocol. These dissimilarities are defined between 0 and 1: when the dissimilarity is close to 0, it means that the patient is following the protocol. In the case where the dissimilarity equals 1, the protocol is not followed at all.

###### Administration dissimilarities

At the level of anti-cancer drug administration, differences can occur in terms of the relative time since the start of the cycle and/or in terms of dosage.

Dose dissimilarity is defined as follows:

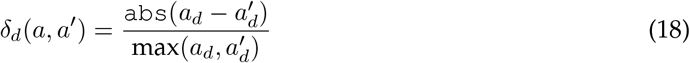

where *a*_*d*_ is the drug dose.

The notation `abs` stands for absolute value to avoid any confusion with the notation |*S*|, which represents the cardinality of a set. According to this definition, *δ*_*d*_(*a, a*^*′*^) is a real number between 0 and

1. When both doses are equal, the dissimilarity is zero. As the administered dose diverges from the theoretical dose, the dissimilarity increases, reaching 1 when the dose is either omitted or added. This aligns with our definition of dissimilarity.

Administration day dissimilarity *δ*_*t*_(*a, a*^*′*^) is defined as follows:

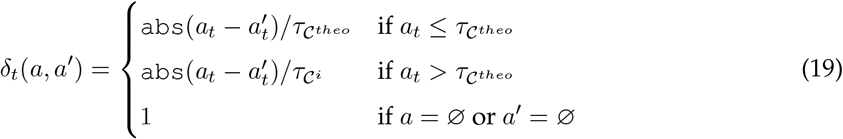

where 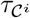 and 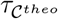 are respectively the durations of the cycles of administrations *a* and *a*^*′*^.

The administration day dissimilarity cannot be normalised in the same way as dose dissimilarity. It cannot be normalised by the maximum between the theoretical administration day and the followed administration day. Indeed, such normalisation would result in higher dissimilarity when administration delays occur at the beginning of the cycle rather than at the end. Thus, when the administration day (*a*_*t*_) is less than the theoretical cycle duration 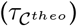, the dissimilarity is normalised by the theoretical cycle duration. When the administration day exceeds the theoretical cycle duration, it is normalised by the followed cycle duration 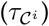. In this way, the administration day dissimilarity is between 0 and 1.

The dissimilarity *δ*_adm_ between two administrations, or between an administration and the absence of administration (if *a* or *a*^*′*^ = Ø), is defined as a weighted sum of the time and dose dissimilarities.

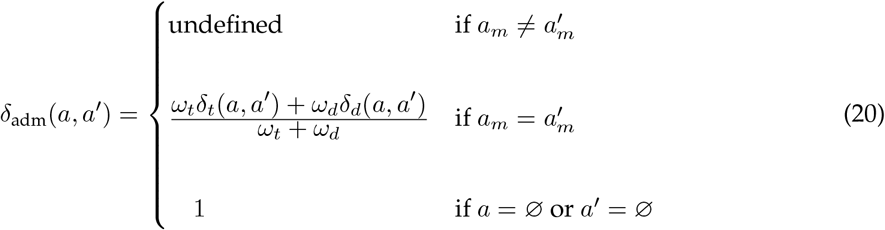

where *ω*_*t*_ and *ω*_*d*_ respectively parameterise the weights associated with time dissimilarity *δ*_*t*_ and dose dissimilarity *δ*_*d*_. The weights are defined in the interval [0, 1] and cannot both be zero.

###### Drug dissimilarity within a cycle

For a given anti-cancer drug *m*, we define drug dissimilarity as the sum of administration dissimilarities for that drug within a cycle. However, the parameters of drug administrations can vary within a cycle. For the same drug *a*_*m*_, there might be different time (*a*_*t*_) and dose (*a*_*d*_) parameters. Therefore, it is necessary to match the theoretical and actual administrations.

For example, in a theoretical cycle, a drug *m* might be scheduled for three different doses on three different days. We define an alignment algorithm to pair the closest matching administrations between the theoretical and actual cycles.

We define *𝒜*_*m*_ as the aligned set of theoretical/actual administration pairs by matching the closest administrations between the theoretical and followed cycles:

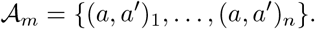

The cardinality of *𝒜*_*m*_ is the maximum number of administrations from either the theoretical or actual cycle: |*𝒜*_*m*_| = max(|*𝒜*^*i*^|, |*𝒜*^*theo*^|). If the number of administrations differs between the two cycles, it means an administration is missing (most often from the actual cycle). In that case, the pair (*a, a*^*′*^)_*i*_ will be represented as (*a*, Ø)_*i*_ or (Ø, *a*^*′*^)_*i*_.

The pseudo-code of the algorithm is presented below.

**Algorithm 1.**
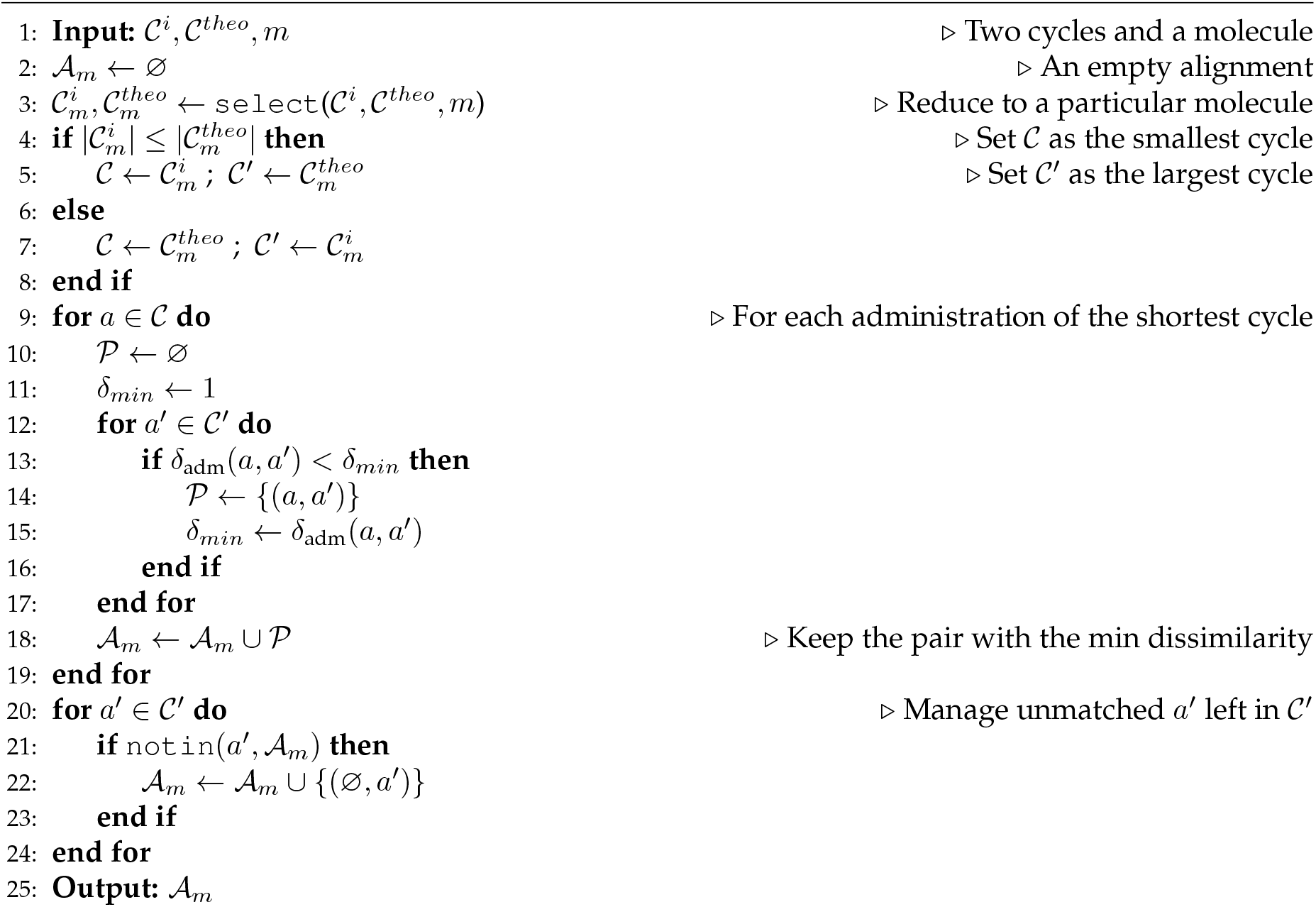
Alignment algorithm

We define the dissimilarity between the theoretical and actual cycles for a drug *m* (a pair of molecule and administration mode) as the average of the administration dissimilarities for that drug:

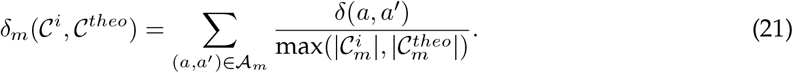

###### Intra and inter cycle dissimilarities

To calculate the intra-cycle dissimilarity, we sum the dissimilarities of the different drugs of the cycle. For each drug *m* (molecule, mode), there can be a drug dissimilarity *δ*_*m*_ within the cycle.

The intra-cycle dissimilarity *δ*_intra_ is defined as:

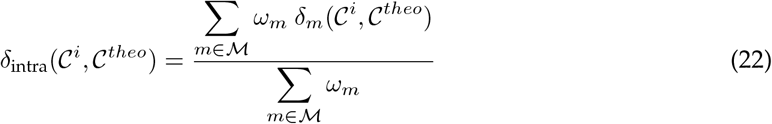

In this article, the weights assigned to each drug are equal, meaning each drug is given the same importance. In this case, the intra-cycle dissimilarity is written as:

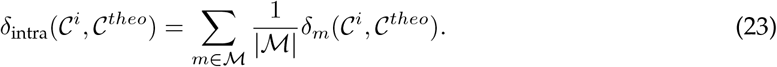

We now introduce inter-cycle dissimilarity, which measures the time gap between two cycles, also called the “inter-cure.” Previously, we considered the timing of drug administrations in relation to the start of each cycle. Now, we will account for differences caused by the time gap between cycles. To explain this dissimilarity more concretely, let’s consider an example. After completing four chemotherapy cycles in December, it is decided to delay the start of the fifth cycle until after the Christmas holidays, allowing the patient to spend time with family. As a result, the fifth cycle begins later than its planned start date.

A simple way to define inter-cycle dissimilarity is to measure the difference between the actual start date of cycle *i* and its theoretical start date. However, this method has a drawback: if cycle 5 is delayed, it will cause delays in all the following cycles as well.

To avoid this issue, we define the dissimilarity for cycle *i* so that it does not depend on delays in earlier cycles. Instead, the delay for cycle *i* is based only on the longer inter-cure period between cycle *i −* 1 and cycle *i*.

The inter-cycle dissimilarity is defined as:

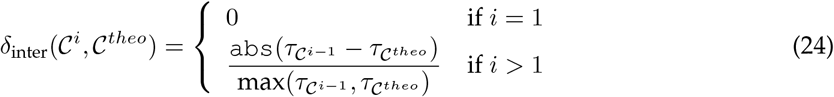

where 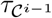 and 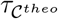 are the durations of cycle *i −* 1 and the theoretical cycle, respectively.

The inter-cycle dissimilarity reflects the delay of cycle *i* due to the longer inter-cure duration of cycle *i −* 1 compared to the theoretical inter-cure. In this way, the delay in cycle *i* does not affect the subsequent cycles (*j > i*).

###### From cycle to line dissimilarities

The dissimilarity between two cycles is a normalised weighted sum of the two intra and inter cycle dimensions:

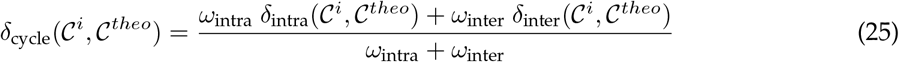

Finally, the line dissimilarity, *δ*_line_, is defined as the cumulative dissimilarity at the last cycle of a line of treatment, as follow:

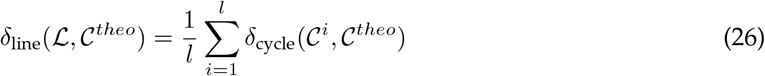

where *l* is the total number of cycles in line *L* (or the last cycle number of the line).

##### A.1.3 Dissimilarity tree

ProtoDrift’s methodology can be visualised as a hierarchical tree, showcasing the relationship between various levels of dissimilarities. This tree structure captures the essence of physicians’ decisions at different treatment stages and time frames, providing a comprehensive view of the treatment deviations

#### A.2 ProtoDrift weights optimisation

##### Dissmilarity reformulations

We provide here the proof of the reformulations of *δ*_adm_ and *δ*_cycle_ as stated in 9 and 10. Given the definition of *α*, we can express *ω*_*d*_ as a function of *ω*_*t*_ and *α*:

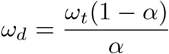

which allows us to write *δ*_adm_ as

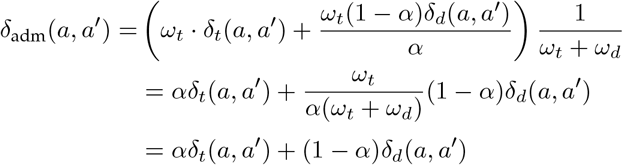

which depends only on *α*. The same reasoning can be applied to express *δ*_cycle_ using *β*.

*δ*_**line**_(*α, β* = 1) **is constant**

For all values of *α, δ*_line_(*α, β* = 1) is a constant. Since the models only vary with the explanatory variable *δ*_line_(*α, β*), and *δ*_line_(*α, β* = 1) is constant, the prediction scores based on this value are also constant.

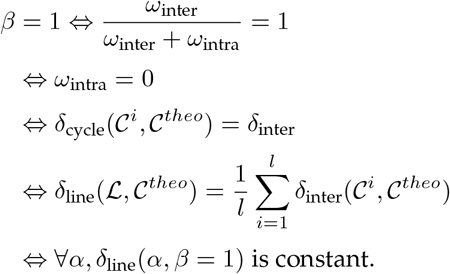

###### Details on the predictive models

The effectiveness of ProtoDrift and its optimised versions against the standard RDI is evaluated using the regression models defined in 11 and 12, and replacing the explanatory variable by ADRDI.

Note that ADRDI quantity is taken at the end of a treatment line, to be comparable with *δ*_line_ dissimilarity.

The three model modalities are summarised in tables A.2 and A.2.

**Table 3:** Logistic regression model formulations.

| Explanatory variable | Model formulation |
| --- | --- |
| ADRFDI | $\text{logit}(P(Y = 1)) = \theta_0 + \theta_1 \text{ADRFDI} + \theta_2 \text{age} + \theta_3 \text{sex}$ |
| $\delta_{\text{line}}(\mathcal{L}, \mathcal{C}^{\text{theo}})_{\alpha=\frac{1}{2}, \beta=\frac{1}{2}}$ | $\text{logit}(P(Y = 1)) = \theta_0 + \theta_1 \delta_{\text{line}}(\mathcal{L}, \mathcal{C}^{\text{theo}})_{\alpha=\frac{1}{2}, \beta=\frac{1}{2}} + \theta_2 \text{age} + \theta_3 \text{sex}$ |
| $\text{argmax}_{\alpha, \beta} \text{AUC}(\delta_{\text{line}}(\mathcal{L}, \mathcal{C}^{\text{theo}})_{\alpha, \beta})$ | $\text{logit}(P(Y = 1)) = \theta_0 + \theta_1 \text{argmax}_{\alpha, \beta} \text{AUC}(\delta_{\text{line}}(\mathcal{L}, \mathcal{C}^{\text{theo}})_{\alpha, \beta}) + \theta_2 \text{age} + \theta_3 \text{sex}$ |

**Table 4:**
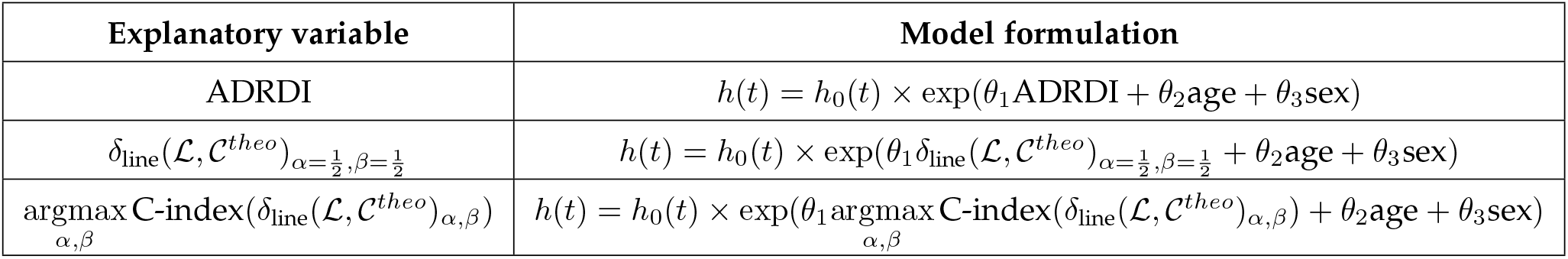
Cox regression model formulations.

### B Results

#### B.1 Study profile

From the patients available in the chemotharapy prescription and administration software (14,662 at HEGP, 33,725 at UHB) we first excluded those for whom it was impossible to compute ProtoDrift or perform a survival analysis due to data quality issues or inconsistencies (2,944 at HEGP, 11,428 at UHB).

Specifically, we removed patients with missing values that prevented us from calculating a normalised administration dose (155 at HEGP, 3 at UHB). Thus, patients on carboplatin regimens were excluded if their creatinine values were missing, as the carboplatin dose is calculated using this data (54 at HEGP, 0 at UHB). We also excluded patients on regimens involving drugs administered in mg/kg if weight data was missing (101 at HEGP, 3 at UHB).

Additionally, patients with inconsistencies in their administration dates (*i*.*e*. doses recorded outside the reported treatment period or after the date of death) were removed (57 at HEGP, 0 at UHB).

We found a number of administrated protocols for which we were unable to find the corresponding theoretical protocol. Patients associated only with these unrecognised protocols were excluded (1,866 at HEGP, 2,949 at UHB).

We also removed patients whose recorded doses were all null (73 at HEGP, 326 at UHB).

We removed patients who followed a regimen with oral administrations, based on the recommendation from pharmacists who noted that oral administrations were not reliably entered in the software (368 at HEGP, 630 at UHB).

Patients with cycles administered to two distinct locations were also removed (117 at HEGP, 594 at UHB).

Finally, patients with missing gender or birth date, were excluded as these data are needed to perform survival analysis (132 at HEGP, 0 at UHB).

Secondly, we excluded patients treated for tumours with fewer than 400 cases (1,956 at HEGP, 4,744 at UHB). At HEGP, patients treated for vulva tumour (517 cases) were removed due to uncertainties regarding the specific conditions referenced in these entries. Additionally, we excluded patients not treated for cancer-related pathologies; this included 1,662 patients at UHB treated for various pathologies and infectious diseases, and patients treated for autoimmune and inflammatory diseases (458 at HEGP and 3,870 at UHB).

#### B.2 Naive ProtoDrift results

For each group, the three Cox indicators, namely Cox regression significance, C-index and LogRank test significance were computed for both NP and ADRDI. Results are summarised in Appendix figures 10 that counts groups where we observe either a gain, loss or balanced result in predicting 5-year overall survival using NP instead of ADRDI.

We observe that, in general, NP outperforms ADRDI. Specifically, 12 out of 18 groups of the HEGP (67%) and 15 out of 20 groups of the UHB (75%) either have comparative or better results with NP. 9 out of 18 groups at HEGP (50%) and 10 out of 20 groups at UHB (50%) have better results for at least one indicator, and better or similar results for the two others. Appendix figure 11a provides the details of those gains per group and per hospital. Over the two hospitals, we observe that it is beneficial to use NP over ADRDI, with at least one better indicator and no loss for Respiratory/Thoracic, Colon, Lymphoid/Haematologic, Pancreas/Biliary Tract, Ovary and Melanoma groups at first line; and for Colon, Bladder/Urothelial, ENT, Ovary, Stomach, Neurology and Melanoma groups at second line.

Notably, four groups at HEGP (22%) and five at UHB (25%) show gains across all the three dimensions (HEGP: Respiratory/Thoracic, Colon and Ovary at first line, ENT at second line; UHB: Colon, ENT, Melanoma first line, Colon and Melanoma second line).

Inversely, ADRDI is better than NP (at least one loss and no gain) in seven groups (18%), which are HEGP ENT and UHB Liver, Bladder/Urothelial, Neurology and Myeloma at first line; HEGP Pancreas/Biliary Tract and UHB Myeloma at second line.

These results suggests that ProtoDrift, even without optimisation, offers a more nuanced capture of treatment adherence compared to ADRDI.

Complete results of the comparative analysis between NP and ADRDI, including performance metrics from Cox and logistic regression models predicting 5-year overall survival across both hospitals, are detailed in Appendix B.3.

#### B.3 Comparative analysis detail results

This annex contains detailed results from our comparative analysis of ProtoDrift against RDI, conducted across two hospitals and focusing on 5-year survival predictions. The results are organised into three sections, corresponding to Cox and logistic regressions across both hospitals.

Each section starts with a table summarising the three performance indicators: significance of regression outcomes, prediction accuracy scores, and the number of significant survival time differences between quartile pairs. The tables use color coding: orange for regression significance, blue for prediction scores, and pink for discriminative power, with significant results (p-values less than 0.05) highlighted in bold.

The second table in each section provides details on the discriminative power analysis, listing the number of significant LogRank test pairs and the survival time differences. These differences are calculated using the median survival times or, for long survival time, the area under the Kaplan-Meier curves calculated through the Restricted Mean Survival Time (RMST).

Results for 3-year survival predictions are available upon request. These tables ensure transparency 30 in presenting our findings.

##### B.3.1 Cox regression - 5 years overall survival

###### HEGP

|  |  | Cox regression fit results |  | Cox regression prediction results |  | Survival time differences |  |
| --- | --- | --- | --- | --- | --- | --- | --- |
| method | Line | $\theta_1$ | p-value | | C-index | Number of significant LogRank tests between quartile pairs | |
| Colon |  |  |  |  |  |  |  |
| OP | 1 | 1.297 | 0.029 |  | 0.521 |  | 3 |
| NP | 1 | 1.682 | 0.044 |  | 0.518 |  | 1 |
| ADRDI | 1 | -0.263 | 0.304 |  | 0.504 |  | 0 |
| OP | 2 | 0.44 | 0.305 |  | 0.493 |  | 0 |
| NP | 2 | 0.102 | 0.447 |  | 0.489 |  | 2 |
| ADRDI | 2 | 0.199 | 0.408 |  | 0.491 |  | 0 |
| Stomach |  |  |  |  |  |  |  |
| OP | 1 | 1.157 | 0.159 |  | 0.533 |  | 0 |
| NP | 1 | 0.697 | 0.387 |  | 0.521 |  | 0 |
| ADRDI | 1 | 0.144 | 0.44 |  | 0.518 |  | 0 |
| OP | 2 | 1.243 | 0.268 |  | 0.511 |  | 0 |
| NP | 2 | 1.558 | 0.31 |  | 0.506 |  | 0 |
| ADRDI | 2 | -0.126 | 0.425 |  | 0.5 |  | 0 |
| Lymph/Hemat |  |  |  |  |  |  |  |
| OP | 1 | 1.351 | 0.101 |  | 0.588 |  | 3 |
| NP | 1 | 1.647 | 0.15 |  | 0.586 |  | 2 |
| ADRDI | 1 | -0.515 | 0.256 |  | 0.581 |  | 1 |
| OP | 2 | -1.358 | 0.234 |  | 0.522 |  | 1 |
| NP | 2 | -1.323 | 0.297 |  | 0.521 |  | 0 |
| ADRDI | 2 | 0.477 | 0.36 |  | 0.52 |  | 0 |
| ENT |  |  |  |  |  |  |  |
| OP | 1 | 0.384 | 0.412 |  | 0.519 |  | 0 |
| NP | 1 | 0.203 | 0.422 |  | 0.518 |  | 0 |
| ADRDI | 1 | -0.526 | 0.129 |  | 0.527 |  | 3 |
| OP | 2 | 1.099 | 0.007 |  | 0.538 |  | 3 |
| NP | 2 | 1.746 | 0.02 |  | 0.53 |  | 3 |
| ADRDI | 2 | -0.659 | 0.106 |  | 0.518 |  | 2 |
| Ovary |  |  |  |  |  |  |  |
| OP | 1 | 1.716 | 0.019 |  | 0.535 |  | 4 |
| NP | 1 | 2.759 | 0.038 |  | 0.524 |  | 3 |
| ADRDI | 1 | -0.441 | 0.357 |  | 0.485 |  | 0 |
| OP | 2 | 1.634 | 0.057 |  | 0.544 |  | 4 |
| NP | 2 | 2.192 | 0.121 |  | 0.525 |  | 2 |
| ADRDI | 2 | -0.73 | 0.276 |  | 0.504 |  | 2 |
| Pancreas/Bil tract |  |  |  |  |  |  |  |
| OP | 1 | 0.931 | 0.051 |  | 0.561 |  | 0 |
| NP | 1 | 0.84 | 0.212 |  | 0.548 |  | 3 |
| ADRDI | 1 | -0.166 | 0.408 |  | 0.541 |  | 0 |
| OP | 2 | -1.7 | 0.077 |  | 0.554 |  | 3 |
| NP | 2 | 0.748 | 0.32 |  | 0.529 |  | 0 |
| ADRDI | 2 | -0.31 | 0.346 |  | 0.527 |  | 1 |
| Respir/Thorac |  |  |  |  |  |  |  |
| OP | 1 | 1.101 | 0 |  | 0.551 |  | 6 |
| NP | 1 | 1.442 | 0.006 |  | 0.538 |  | 4 |
| ADRDI | 1 | -0.283 | 0.234 |  | 0.519 |  | 0 |
| OP | 2 | 0.713 | 0.072 |  | 0.527 |  | 2 |
| NP | 2 | 0.947 | 0.149 |  | 0.523 |  | 1 |
| ADRDI | 2 | -0.255 | 0.348 |  | 0.515 |  | 2 |
| Breast |  |  |  |  |  |  |  |
| OP | 1 | 5.032 | 0 |  | 0.655 |  | 6 |
| NP | 1 | 4.581 | 0 |  | 0.653 |  | 6 |
| ADRDI | 1 | -2.567 | 0 |  | 0.663 |  | 5 |
| OP | 2 | 3.942 | 0 |  | 0.595 |  | 5 |
| NP | 2 | 3.92 | 0 |  | 0.594 |  | 4 |
| ADRDI | 2 | -1.806 | 0.003 |  | 0.57 |  | 5 |
| Bladder/Uroth |  |  |  |  |  |  |  |
| OP | 1 | 1.222 | 0.01 |  | 0.552 |  | 4 |
| NP | 1 | 1.167 | 0.173 |  | 0.53 |  | 2 |
| ADRDI | 1 | -0.17 | 0.407 |  | 0.519 |  | 3 |
| OP | 2 | 1.982 | 0.047 |  | 0.553 |  | 4 |
| NP | 2 | 2.881 | 0.078 |  | 0.536 |  | 5 |
| ADRDI | 2 | -0.375 | 0.371 |  | 0.498 |  | 2 |

Results of three performance indicators for HEGP patient groups

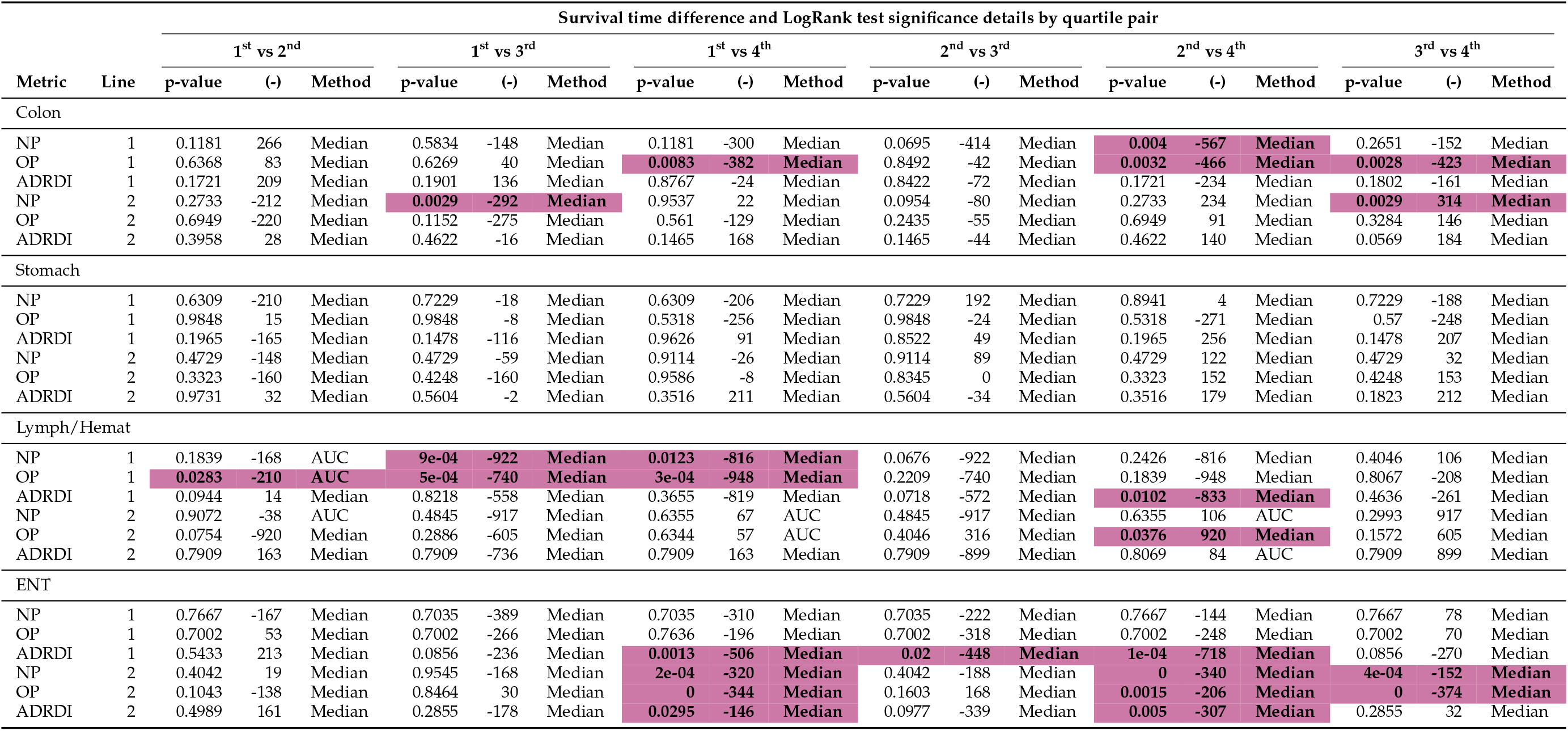

Detailed discriminative power results for HEGP patient groups (1/3)

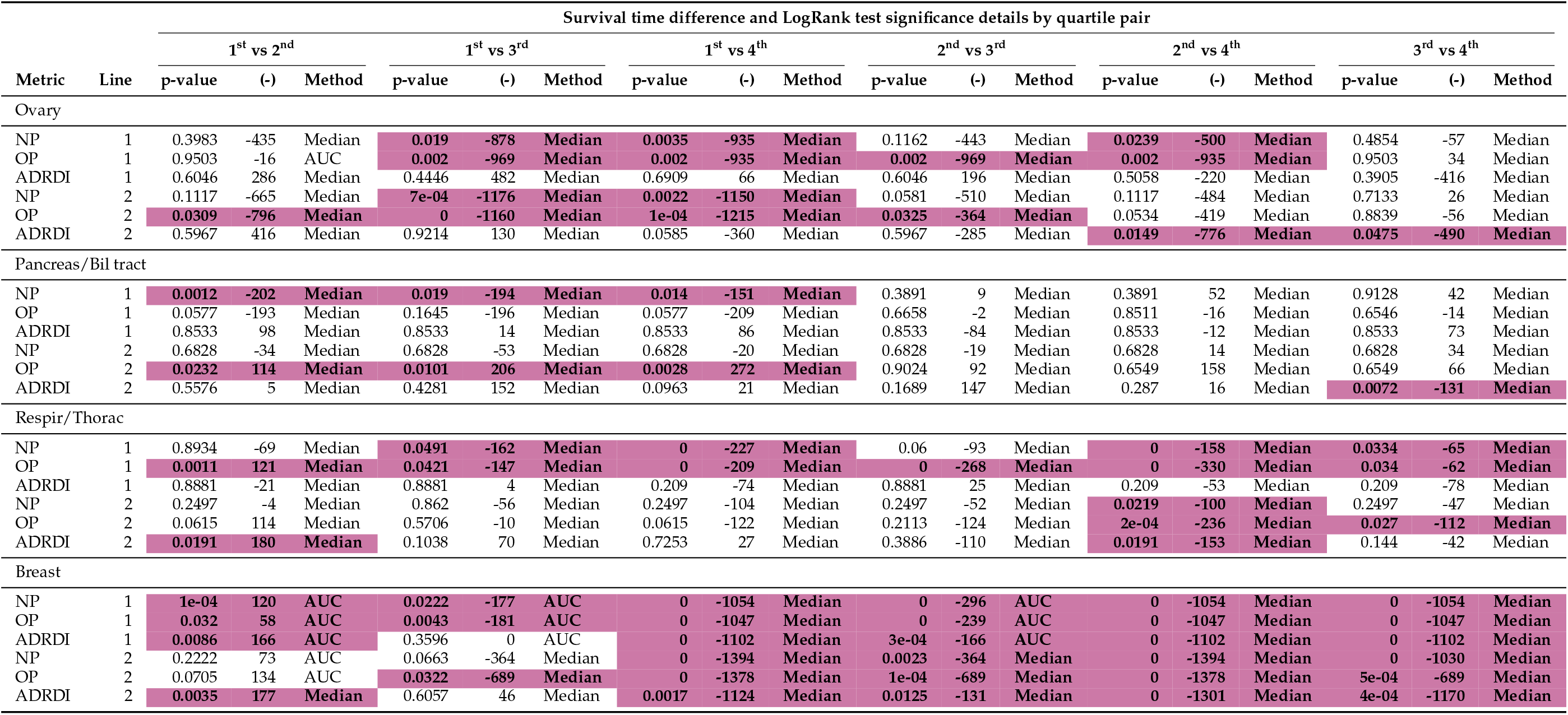

| Survival time difference and LogRank test significance details by quartile pair |  |  |  |  |  |  |  |  |  |  |  |  |  |  |  |  |  |  |  |
| --- | --- | --- | --- | --- | --- | --- | --- | --- | --- | --- | --- | --- | --- | --- | --- | --- | --- | --- | --- |
| Metric | Line | 1<sup>st</sup> vs 2<sup>nd</sup> |  |  | 1<sup>st</sup> vs 3<sup>rd</sup> |  |  | 1<sup>st</sup> vs 4<sup>th</sup> |  |  | 2<sup>nd</sup> vs 3<sup>rd</sup> |  |  | 2<sup>nd</sup> vs 4<sup>th</sup> |  |  | 3<sup>rd</sup> vs 4<sup>th</sup> |  |  |
|  |  | p-value | (-) | Method | p-value | (-) | Method | p-value | (-) | Method | p-value | (-) | Method | p-value | (-) | Method | p-value | (-) | Method |
| Ovary |  |  |  |  |  |  |  |  |  |  |  |  |  |  |  |  |  |  |  |
| NP | 1 | 0.3983 | -435 | Median | 0.019 | -878 | Median | 0.0035 | -935 | Median | 0.1162 | -443 | Median | 0.0239 | -500 | Median | 0.4854 | -57 | Median |
| OP | 1 | 0.9503 | -16 | AUC | 0.002 | -969 | Median | 0.002 | -935 | Median | 0.002 | -969 | Median | 0.002 | -935 | Median | 0.9503 | 34 | Median |
| ADRD | 1 | 0.6046 | 286 | Median | 0.4446 | 482 | Median | 0.6909 | 66 | Median | 0.6046 | 196 | Median | 0.5058 | -220 | Median | 0.3905 | -416 | Median |
| NP | 2 | 0.1117 | -665 | Median | 7e-04 | -1176 | Median | 0.0022 | -1150 | Median | 0.0581 | -510 | Median | 0.1117 | -484 | Median | 0.7133 | 26 | Median |
| OP | 2 | 0.0309 | -796 | Median | 0 | -1160 | Median | 1e-04 | -1215 | Median | 0.0325 | -364 | Median | 0.0534 | -419 | Median | 0.8839 | -56 | Median |
| ADRD | 2 | 0.5967 | 416 | Median | 0.9214 | 130 | Median | 0.0585 | -360 | Median | 0.5967 | -285 | Median | 0.0149 | -776 | Median | 0.0475 | -490 | Median |
| Pancreas/Bil tract |  |  |  |  |  |  |  |  |  |  |  |  |  |  |  |  |  |  |  |
| NP | 1 | 0.0012 | -202 | Median | 0.019 | -194 | Median | 0.014 | -151 | Median | 0.3891 | 9 | Median | 0.3891 | 52 | Median | 0.9128 | 42 | Median |
| OP | 1 | 0.0577 | -193 | Median | 0.1645 | -196 | Median | 0.0577 | -209 | Median | 0.6658 | -2 | Median | 0.8511 | -16 | Median | 0.6546 | -14 | Median |
| ADRD | 1 | 0.8533 | 98 | Median | 0.8533 | 14 | Median | 0.8533 | 86 | Median | 0.8533 | -84 | Median | 0.8533 | -12 | Median | 0.8533 | 73 | Median |
| NP | 2 | 0.6828 | -34 | Median | 0.6828 | -53 | Median | 0.6828 | -20 | Median | 0.6828 | -19 | Median | 0.6828 | 14 | Median | 0.6828 | 34 | Median |
| OP | 2 | 0.0232 | 114 | Median | 0.0101 | 206 | Median | 0.0028 | 272 | Median | 0.9024 | 92 | Median | 0.6549 | 158 | Median | 0.6549 | 66 | Median |
| ADRD | 2 | 0.5576 | 5 | Median | 0.4281 | 152 | Median | 0.0963 | 21 | Median | 0.1689 | 147 | Median | 0.287 | 16 | Median | 0.0072 | -131 | Median |
| Respir/Thorac |  |  |  |  |  |  |  |  |  |  |  |  |  |  |  |  |  |  |  |
| NP | 1 | 0.8934 | -69 | Median | 0.0491 | -162 | Median | 0 | -227 | Median | 0.06 | -93 | Median | 0 | -158 | Median | 0.0334 | -65 | Median |
| OP | 1 | 0.0011 | 121 | Median | 0.0421 | -147 | Median | 0 | -209 | Median | 0 | -268 | Median | 0 | -330 | Median | 0.034 | -62 | Median |
| ADRD | 1 | 0.8881 | -21 | Median | 0.8881 | 4 | Median | 0.209 | -74 | Median | 0.8881 | 25 | Median | 0.209 | -53 | Median | 0.209 | -78 | Median |
| NP | 2 | 0.2497 | -4 | Median | 0.862 | -56 | Median | 0.2497 | -104 | Median | 0.2497 | -52 | Median | 0.0219 | -100 | Median | 0.2497 | -47 | Median |
| OP | 2 | 0.0615 | 114 | Median | 0.5706 | -10 | Median | 0.0615 | -122 | Median | 0.2113 | -124 | Median | 2e-04 | -236 | Median | 0.027 | -112 | Median |
| ADRD | 2 | 0.0191 | 180 | Median | 0.1038 | 70 | Median | 0.7253 | 27 | Median | 0.3886 | -110 | Median | 0.0191 | -153 | Median | 0.144 | -42 | Median |
| Breast |  |  |  |  |  |  |  |  |  |  |  |  |  |  |  |  |  |  |  |
| NP | 1 | 1e-04 | 120 | AUC | 0.0222 | -177 | AUC | 0 | -1054 | Median | 0 | -296 | AUC | 0 | -1054 | Median | 0 | -1054 | Median |
| OP | 1 | 0.032 | 58 | AUC | 0.0043 | -181 | AUC | 0 | -1047 | Median | 0 | -239 | AUC | 0 | -1047 | Median | 0 | -1047 | Median |
| ADRD | 1 | 0.0086 | 166 | AUC | 0.3596 | 0 | AUC | 0 | -1102 | Median | 3e-04 | -166 | AUC | 0 | -1102 | Median | 0 | -1102 | Median |
| NP | 2 | 0.2222 | 73 | AUC | 0.0663 | -364 | Median | 0 | -1394 | Median | 0.0023 | -364 | Median | 0 | -1394 | Median | 0 | -1030 | Median |
| OP | 2 | 0.0705 | 134 | AUC | 0.0322 | -689 | Median | 0 | -1378 | Median | 1e-04 | -689 | Median | 0 | -1378 | Median | 5e-04 | -689 | Median |
| ADRD | 2 | 0.0035 | 177 | Median | 0.6057 | 46 | Median | 0.0017 | -1124 | Median | 0.0125 | -131 | Median | 0 | -1301 | Median | 4e-04 | -1170 | Median |

Detailed discriminative power results for HEGP patient groups (2/3)

| Survival time difference and LogRank test significance details by quartile pair |  |  |  |  |  |  |  |  |  |  |  |  |  |  |  |  |  |  |  |
| --- | --- | --- | --- | --- | --- | --- | --- | --- | --- | --- | --- | --- | --- | --- | --- | --- | --- | --- | --- |
| Metric | Line | 1<sup>st</sup> vs 2<sup>nd</sup> |  |  | 1<sup>st</sup> vs 3<sup>rd</sup> |  |  | 1<sup>st</sup> vs 4<sup>th</sup> |  |  | 2<sup>nd</sup> vs 3<sup>rd</sup> |  |  | 2<sup>nd</sup> vs 4<sup>th</sup> |  |  | 3<sup>rd</sup> vs 4<sup>th</sup> |  |  |
|  |  | p-value | (-) | Method | p-value | (-) | Method | p-value | (-) | Method | p-value | (-) | Method | p-value | (-) | Method | p-value | (-) | Method |
| Bladder/Uroth |  |  |  |  |  |  |  |  |  |  |  |  |  |  |  |  |  |  |  |
| NP | 1 | 0.1119 | -300 | Median | 0.8734 | 162 | Median | 0.0101 | -523 | Median | 0.1325 | 462 | Median | 0.2259 | -223 | Median | 0.0101 | -685 | Median |
| OP | 1 | 0.0571 | 843 | Median | 0.0452 | -353 | Median | 4e-04 | -570 | Median | 2e-04 | -1196 | Median | 0 | -1413 | Median | 0.1414 | -217 | Median |
| ADRFI | 1 | 0.707 | -64 | Median | 0.0061 | -492 | Median | 0.707 | 138 | Median | 0.0077 | -428 | Median | 0.9286 | 202 | Median | 0.0077 | 630 | Median |
| NP | 2 | 0.0363 | -370 | Median | 0.0363 | -344 | Median | 1e-04 | -576 | Median | 0.9938 | 26 | Median | 0.0363 | -206 | Median | 0.0363 | -232 | Median |
| OP | 2 | 0.9468 | -34 | Median | 0.0026 | -418 | Median | 0.001 | -492 | Median | 0.0022 | -383 | Median | 9e-04 | -457 | Median | 0.4839 | -74 | Median |
| ADRFI | 2 | 0.8227 | -124 | Median | 0.0432 | -142 | Median | 0.4786 | -218 | Median | 0.0474 | -19 | Median | 0.4786 | -94 | Median | 0.2391 | -75 | Median |

Detailed discriminative power results for HEGP patient groups (3/3)

**Table 10:**
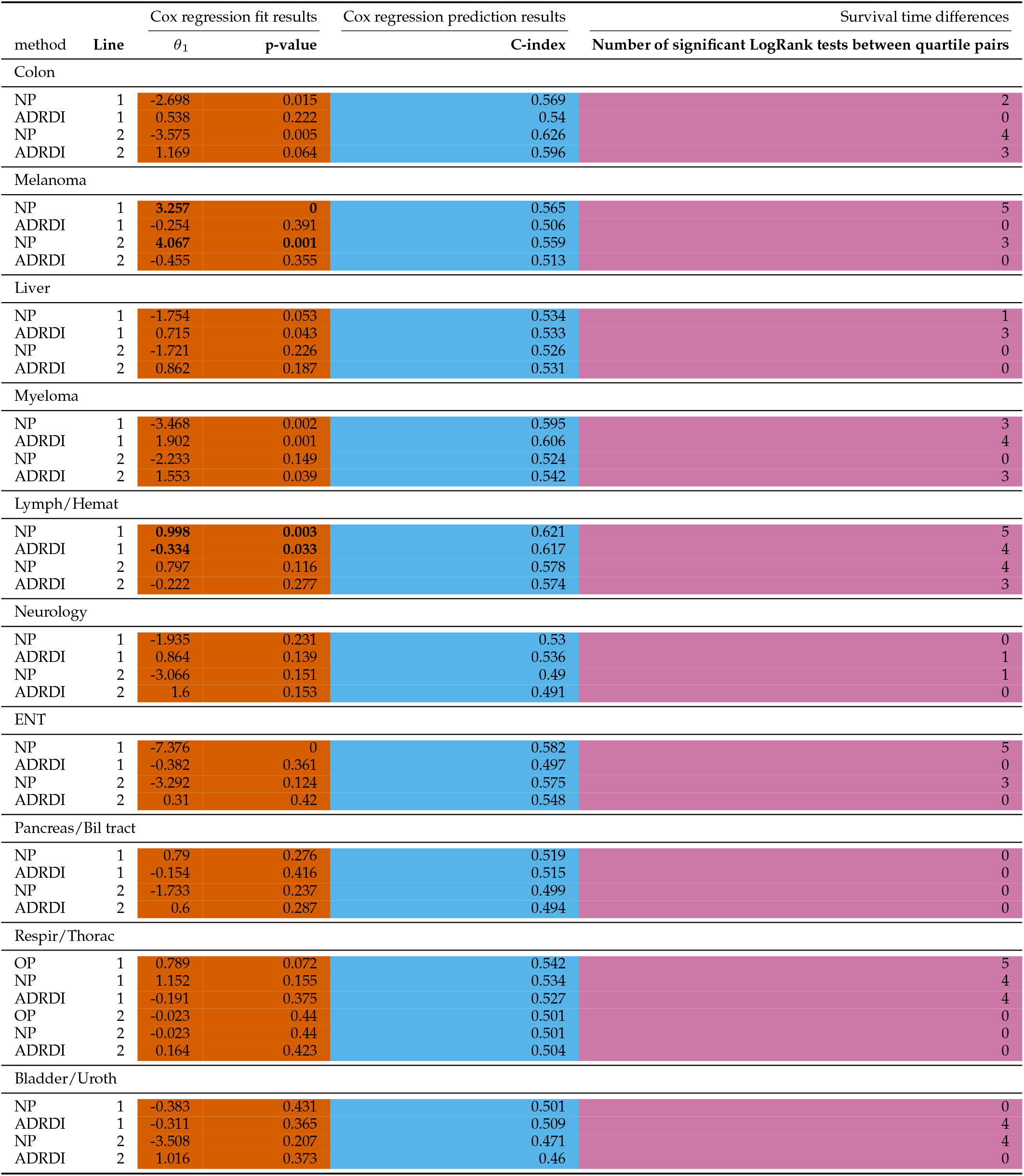
Results of three performance indicators for UHB patient groups.

###### UHB

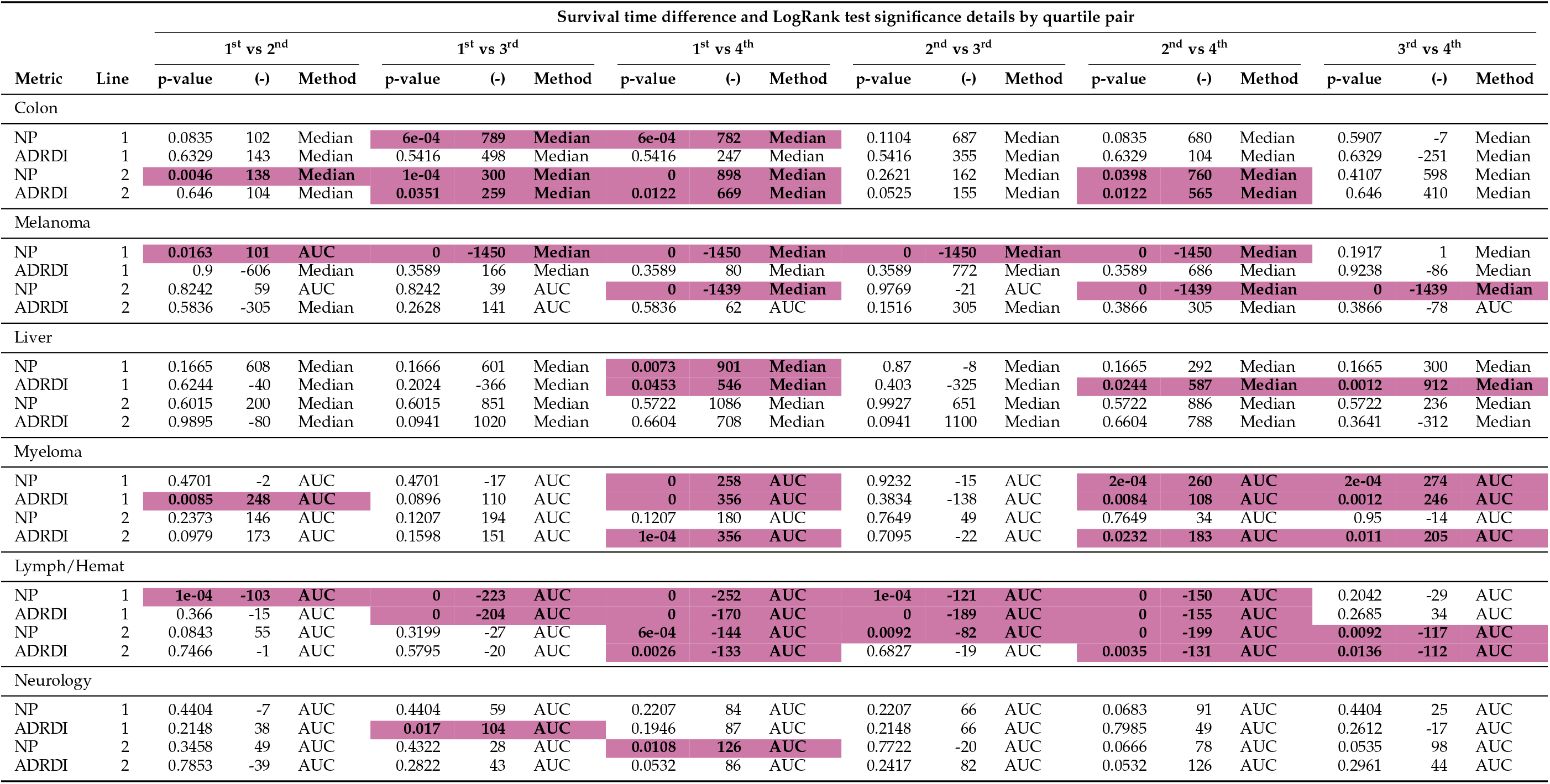

Detailed discriminative power results for UHB patient groups (1/2)

| Survival time difference and LogRank test significance details by quartile pair |  |  |  |  |  |  |  |  |  |  |  |  |  |  |  |  |  |  |  |
| --- | --- | --- | --- | --- | --- | --- | --- | --- | --- | --- | --- | --- | --- | --- | --- | --- | --- | --- | --- |
| Metric | Line | 1<sup>st</sup> vs 2<sup>nd</sup> |  |  | 1<sup>st</sup> vs 3<sup>rd</sup> |  |  | 1<sup>st</sup> vs 4<sup>th</sup> |  |  | 2<sup>nd</sup> vs 3<sup>rd</sup> |  |  | 2<sup>nd</sup> vs 4<sup>th</sup> |  |  | 3<sup>rd</sup> vs 4<sup>th</sup> |  |  |
|  |  | p-value | (-) | Method | p-value | (-) | Method | p-value | (-) | Method | p-value | (-) | Method | p-value | (-) | Method | p-value | (-) | Method |
| ENT |  |  |  |  |  |  |  |  |  |  |  |  |  |  |  |  |  |  |  |
| NP | 1 | 0 | 1443 | Median | 0 | 1443 | Median | 0 | 1443 | Median | 0 | 334 | AUC | 0.005 | 196 | AUC | 0.2009 | -138 | AUC |
| ADRFI | 1 | 0.8001 | -23 | AUC | 0.1125 | -254 | Median | 0.1882 | -176 | AUC | 0.1125 | -254 | Median | 0.1854 | -153 | AUC | 0.8001 | 254 | Median |
| NP | 2 | 0.2274 | 138 | Median | 0.0347 | 980 | Median | 0.001 | 1484 | Median | 0.2274 | 842 | Median | 0.0144 | 1346 | Median | 0.1934 | 504 | Median |
| ADRFI | 2 | 0.3918 | 1013 | Median | 0.5009 | -26 | Median | 0.551 | 299 | Median | 0.0876 | -1039 | Median | 0.551 | -714 | Median | 0.2908 | 325 | Median |
| Pancreas/Bil tract |  |  |  |  |  |  |  |  |  |  |  |  |  |  |  |  |  |  |  |
| NP | 1 | 0.9656 | 15 | Median | 0.3544 | 36 | Median | 0.3544 | -5 | Median | 0.3544 | 21 | Median | 0.3544 | -20 | Median | 0.1046 | -41 | Median |
| ADRFI | 1 | 0.0662 | 148 | Median | 0.0662 | 115 | Median | 0.9505 | 95 | Median | 0.9697 | -34 | Median | 0.0662 | -54 | Median | 0.0662 | -20 | Median |
| NP | 2 | 0.1838 | 160 | Median | 0.3803 | 106 | Median | 0.0705 | 158 | Median | 0.4522 | -54 | Median | 0.3803 | -2 | Median | 0.2045 | 52 | Median |
| ADRFI | 2 | 0.5642 | 138 | Median | 0.4382 | 132 | Median | 0.4382 | 69 | Median | 0.7664 | -6 | Median | 0.7664 | -69 | Median | 0.7664 | -63 | Median |
| Respir/Thorac |  |  |  |  |  |  |  |  |  |  |  |  |  |  |  |  |  |  |  |
| NP | 1 | 0.0848 | -472 | Median | 1e-04 | -670 | Median | 0.4012 | 846 | Median | 0.0242 | -198 | Median | 0.0242 | 1318 | Median | 0 | 1516 | Median |
| OP | 1 | 0.0144 | -697 | Median | 0 | -726 | Median | 0.3368 | 792 | Median | 0.0063 | -28 | Median | 0.0063 | 1488 | Median | 0 | 1517 | Median |
| ADRFI | 1 | 0.0218 | 667 | Median | 0.982 | 200 | Median | 0.0316 | 1100 | Median | 0.0218 | -468 | Median | 0.982 | 432 | Median | 0.0218 | 900 | Median |
| NP | 2 | 0.8153 | -199 | Median | 0.8554 | 138 | Median | 0.9394 | 60 | Median | 0.8554 | 338 | Median | 0.8153 | 258 | Median | 0.8554 | -79 | Median |
| OP | 2 | 0.8153 | -199 | Median | 0.8554 | 138 | Median | 0.9394 | 60 | Median | 0.8554 | 338 | Median | 0.8153 | 258 | Median | 0.8554 | -79 | Median |
| ADRFI | 2 | 0.4824 | 66 | Median | 0.8894 | 128 | Median | 0.2451 | 406 | Median | 0.4824 | 62 | Median | 0.4824 | 340 | Median | 0.2451 | 278 | Median |
| Bladder/Uroth |  |  |  |  |  |  |  |  |  |  |  |  |  |  |  |  |  |  |  |
| NP | 1 | 0.6336 | -96 | AUC | 0.6336 | -50 | AUC | 0.8848 | 21 | AUC | 0.9268 | 46 | AUC | 0.6336 | 117 | AUC | 0.6336 | 71 | AUC |
| ADRFI | 1 | 0.5754 | 26 | AUC | 5e-04 | -335 | AUC | 7e-04 | -296 | AUC | 1e-04 | -361 | AUC | 1e-04 | -322 | AUC | 0.8399 | 40 | AUC |
| NP | 2 | 0.0443 | 461 | AUC | 0.6918 | 45 | AUC | 0.0318 | 548 | AUC | 0.0482 | -416 | AUC | 0.6918 | 87 | AUC | 0.0318 | 503 | AUC |
| ADRFI | 2 | 0.5006 | -159 | AUC | 0.5006 | -249 | AUC | 0.9255 | 66 | AUC | 0.9244 | -90 | AUC | 0.5006 | 225 | AUC | 0.5006 | 315 | AUC |

Detailed discriminative power results for UHB patient groups (2/2)

##### B.3.2 Logistic regression - 5 years overall survival HEGP

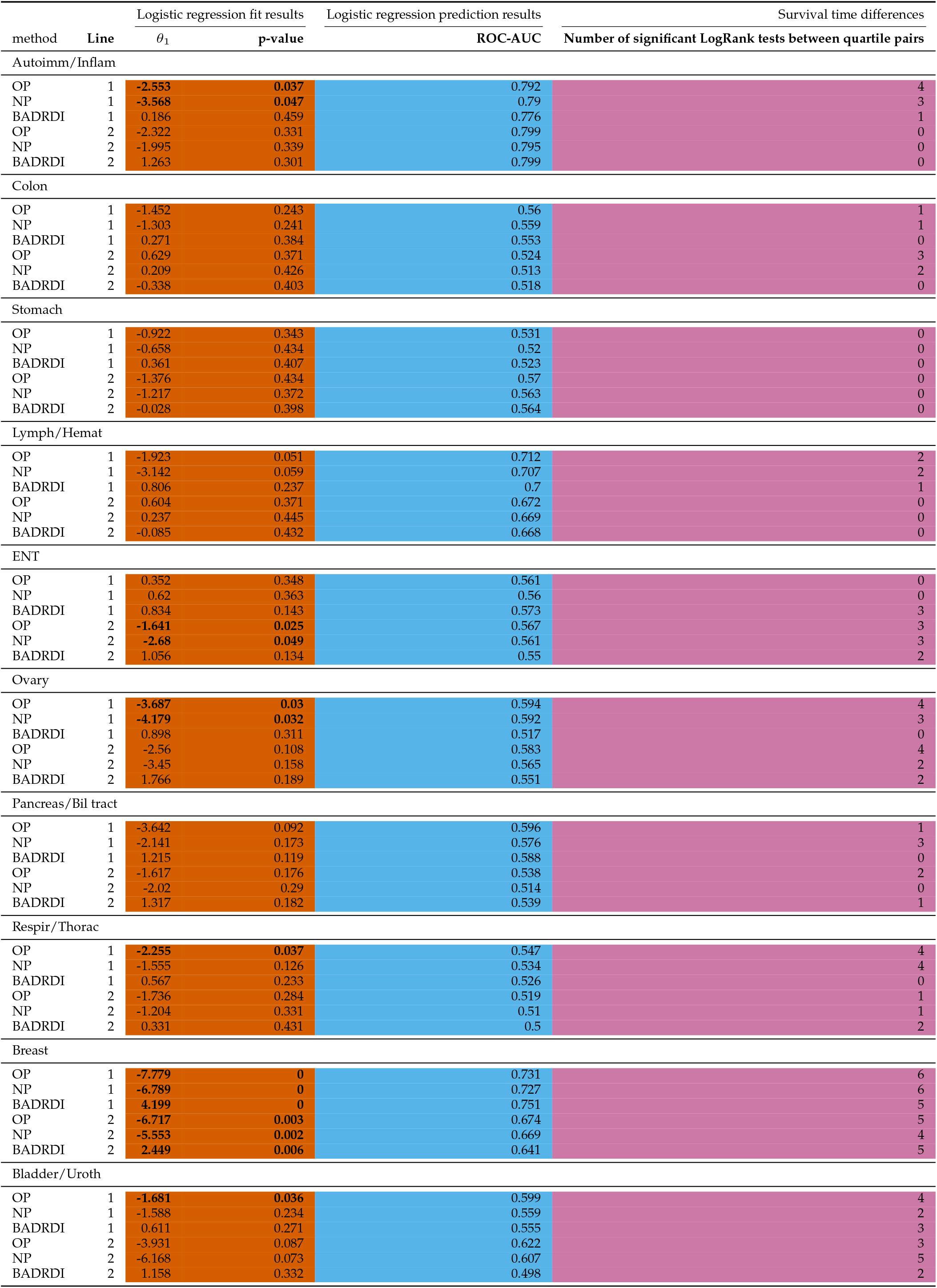

|  |  | Logistic regression fit results |  | Logistic regression prediction results |  | Survival time differences |  |
| --- | --- | --- | --- | --- | --- | --- | --- |
| method | Line | $\theta_1$ | p-value | ROC-AUC | | Number of significant LogRank tests between quartile pairs | |
| Autoimm/Inflam |  |  |  |  |  |  |  |
| OP | 1 | -2.553 | 0.037 | 0.792 |  | 4 |  |
| NP | 1 | -3.568 | 0.047 | 0.79 |  | 3 |  |
| BADRDI | 1 | 0.186 | 0.459 | 0.776 |  | 1 |  |
| OP | 2 | -2.322 | 0.331 | 0.799 |  | 0 |  |
| NP | 2 | -1.995 | 0.339 | 0.795 |  | 0 |  |
| BADRDI | 2 | 1.263 | 0.301 | 0.799 |  | 0 |  |
| Colon |  |  |  |  |  |  |  |
| OP | 1 | -1.452 | 0.243 | 0.56 |  | 1 |  |
| NP | 1 | -1.303 | 0.241 | 0.559 |  | 1 |  |
| BADRDI | 1 | 0.271 | 0.384 | 0.553 |  | 0 |  |
| OP | 2 | 0.629 | 0.371 | 0.524 |  | 3 |  |
| NP | 2 | 0.209 | 0.426 | 0.513 |  | 2 |  |
| BADRDI | 2 | -0.338 | 0.403 | 0.518 |  | 0 |  |
| Stomach |  |  |  |  |  |  |  |
| OP | 1 | -0.922 | 0.343 | 0.531 |  | 0 |  |
| NP | 1 | -0.658 | 0.434 | 0.52 |  | 0 |  |
| BADRDI | 1 | 0.361 | 0.407 | 0.523 |  | 0 |  |
| OP | 2 | -1.376 | 0.434 | 0.57 |  | 0 |  |
| NP | 2 | -1.217 | 0.372 | 0.563 |  | 0 |  |
| BADRDI | 2 | -0.028 | 0.398 | 0.564 |  | 0 |  |
| Lymph/Hemat |  |  |  |  |  |  |  |
| OP | 1 | -1.923 | 0.051 | 0.712 |  | 2 |  |
| NP | 1 | -3.142 | 0.059 | 0.707 |  | 2 |  |
| BADRDI | 1 | 0.806 | 0.237 | 0.7 |  | 1 |  |
| OP | 2 | 0.604 | 0.371 | 0.672 |  | 0 |  |
| NP | 2 | 0.237 | 0.445 | 0.669 |  | 0 |  |
| BADRDI | 2 | -0.085 | 0.432 | 0.668 |  | 0 |  |
| ENT |  |  |  |  |  |  |  |
| OP | 1 | 0.352 | 0.348 | 0.561 |  | 0 |  |
| NP | 1 | 0.62 | 0.363 | 0.56 |  | 0 |  |
| BADRDI | 1 | 0.834 | 0.143 | 0.573 |  | 3 |  |
| OP | 2 | -1.641 | 0.025 | 0.567 |  | 3 |  |
| NP | 2 | -2.68 | 0.049 | 0.561 |  | 3 |  |
| BADRDI | 2 | 1.056 | 0.134 | 0.55 |  | 2 |  |
| Ovary |  |  |  |  |  |  |  |
| OP | 1 | -3.687 | 0.03 | 0.594 |  | 4 |  |
| NP | 1 | -4.179 | 0.032 | 0.592 |  | 3 |  |
| BADRDI | 1 | 0.898 | 0.311 | 0.517 |  | 0 |  |
| OP | 2 | -2.56 | 0.108 | 0.583 |  | 4 |  |
| NP | 2 | -3.45 | 0.158 | 0.565 |  | 2 |  |
| BADRDI | 2 | 1.766 | 0.189 | 0.551 |  | 2 |  |
| Pancreas/Bil tract |  |  |  |  |  |  |  |
| OP | 1 | -3.642 | 0.092 | 0.596 |  | 1 |  |
| NP | 1 | -2.141 | 0.173 | 0.576 |  | 3 |  |
| BADRDI | 1 | 1.215 | 0.119 | 0.588 |  | 0 |  |
| OP | 2 | -1.617 | 0.176 | 0.538 |  | 2 |  |
| NP | 2 | -2.02 | 0.29 | 0.514 |  | 0 |  |
| BADRDI | 2 | 1.317 | 0.182 | 0.539 |  | 1 |  |
| Respir/Thorac |  |  |  |  |  |  |  |
| OP | 1 | -2.255 | 0.037 | 0.547 |  | 4 |  |
| NP | 1 | -1.555 | 0.126 | 0.534 |  | 4 |  |
| BADRDI | 1 | 0.567 | 0.233 | 0.526 |  | 0 |  |
| OP | 2 | -1.736 | 0.284 | 0.519 |  | 1 |  |
| NP | 2 | -1.204 | 0.331 | 0.51 |  | 1 |  |
| BADRDI | 2 | 0.331 | 0.431 | 0.5 |  | 2 |  |
| Breast |  |  |  |  |  |  |  |
| OP | 1 | -7.779 | 0 | 0.731 |  | 6 |  |
| NP | 1 | -6.789 | 0 | 0.727 |  | 6 |  |
| BADRDI | 1 | 4.199 | 0 | 0.751 |  | 5 |  |
| OP | 2 | -6.717 | 0.003 | 0.674 |  | 5 |  |
| NP | 2 | -5.553 | 0.002 | 0.669 |  | 4 |  |
| BADRDI | 2 | 2.449 | 0.006 | 0.641 |  | 5 |  |
| Bladder/Uroth |  |  |  |  |  |  |  |
| OP | 1 | -1.681 | 0.036 | 0.599 |  | 4 |  |
| NP | 1 | -1.588 | 0.234 | 0.559 |  | 2 |  |
| BADRDI | 1 | 0.611 | 0.271 | 0.555 |  | 3 |  |
| OP | 2 | -3.931 | 0.087 | 0.622 |  | 3 |  |
| NP | 2 | -6.168 | 0.073 | 0.607 |  | 5 |  |
| BADRDI | 2 | 1.158 | 0.332 | 0.498 |  | 2 |  |

Results of three performance indicators for H41EGP patient groups using logistic regression

Results of three performance indicators for LIEGP patient groups using logistic regression
| Survival time difference and LogRank test significance details by quartile pair |  |  |  |  |  |  |  |  |  |  |  |  |  |  |  |  |  |  |  |
| --- | --- | --- | --- | --- | --- | --- | --- | --- | --- | --- | --- | --- | --- | --- | --- | --- | --- | --- | --- |
| Metric | Line | 1<sup>st</sup> vs 2<sup>nd</sup> |  |  | 1<sup>st</sup> vs 3<sup>rd</sup> |  |  | 1<sup>st</sup> vs 4<sup>th</sup> |  |  | 2<sup>nd</sup> vs 3<sup>rd</sup> |  |  | 2<sup>nd</sup> vs 4<sup>th</sup> |  |  | 3<sup>rd</sup> vs 4<sup>th</sup> |  |  |
|  |  | p-value | (-) | Method | p-value | (-) | Method | p-value | (-) | Method | p-value | (-) | Method | p-value | (-) | Method | p-value | (-) | Method |
| Colon |  |  |  |  |  |  |  |  |  |  |  |  |  |  |  |  |  |  |  |
| NP | 1 | 0.1181 | 266 | Median | 0.5834 | -148 | Median | 0.1181 | -300 | Median | 0.0695 | -414 | Median | 0.004 | -567 | Median | 0.2651 | -152 | Median |
| OP | 1 | 0.3493 | 236 | Median | 0.6998 | -104 | Median | 0.1246 | -302 | Median | 0.2423 | -341 | Median | 0.0165 | -538 | Median | 0.1928 | -197 | Median |
| ADRFI | 1 | 0.1721 | 209 | Median | 0.1901 | 136 | Median | 0.8767 | -24 | Median | 0.8422 | -72 | Median | 0.1721 | -234 | Median | 0.1802 | -161 | Median |
| NP | 2 | 0.2733 | -212 | Median | 0.0029 | -292 | Median | 0.9537 | 22 | Median | 0.0954 | -80 | Median | 0.2733 | 234 | Median | 0.0029 | 314 | Median |
| OP | 2 | 0.8584 | -23 | Median | 0.0476 | -213 | Median | 0.7687 | 67 | Median | 0.0476 | -190 | Median | 0.783 | 90 | Median | 0.031 | 280 | Median |
| ADRFI | 2 | 0.3958 | 28 | Median | 0.4622 | -16 | Median | 0.1465 | 168 | Median | 0.1465 | -44 | Median | 0.4622 | 140 | Median | 0.0569 | 184 | Median |
| Stomach |  |  |  |  |  |  |  |  |  |  |  |  |  |  |  |  |  |  |  |
| NP | 1 | 0.6309 | -210 | Median | 0.7229 | -18 | Median | 0.6309 | -206 | Median | 0.7229 | 192 | Median | 0.8941 | 4 | Median | 0.7229 | -188 | Median |
| OP | 1 | 0.9848 | 15 | Median | 0.9848 | -8 | Median | 0.5318 | -256 | Median | 0.9848 | -24 | Median | 0.5318 | -271 | Median | 0.57 | -248 | Median |
| ADRFI | 1 | 0.1965 | -165 | Median | 0.1478 | -116 | Median | 0.9626 | 91 | Median | 0.8522 | 49 | Median | 0.1965 | 256 | Median | 0.1478 | 207 | Median |
| NP | 2 | 0.4729 | -148 | Median | 0.4729 | -59 | Median | 0.9114 | -26 | Median | 0.9114 | 89 | Median | 0.4729 | 122 | Median | 0.4729 | 32 | Median |
| OP | 2 | 0.9112 | -23 | Median | 0.9112 | 64 | Median | 0.9112 | 98 | Median | 0.9112 | 86 | Median | 0.9112 | 122 | Median | 0.9112 | 35 | Median |
| ADRFI | 2 | 0.9731 | 32 | Median | 0.5604 | -2 | Median | 0.3516 | 211 | Median | 0.5604 | -34 | Median | 0.3516 | 179 | Median | 0.1823 | 212 | Median |
| Lymph/Hemat |  |  |  |  |  |  |  |  |  |  |  |  |  |  |  |  |  |  |  |
| NP | 1 | 0.1839 | -168 | AUC | 9e-04 | -922 | Median | 0.0123 | -816 | Median | 0.0676 | -922 | Median | 0.2426 | -816 | Median | 0.4046 | 106 | Median |
| OP | 1 | 0.0579 | -173 | AUC | 0.0016 | -686 | Median | 2e-04 | -1080 | Median | 0.192 | -686 | Median | 0.0579 | -1080 | Median | 0.4729 | -394 | Median |
| ADRFI | 1 | 0.0944 | 14 | Median | 0.8218 | -558 | Median | 0.3655 | -819 | Median | 0.0718 | -572 | Median | 0.0102 | -833 | Median | 0.4636 | -261 | Median |
| NP | 2 | 0.9072 | -38 | AUC | 0.4845 | -917 | Median | 0.6355 | 67 | AUC | 0.4845 | -917 | Median | 0.6355 | 106 | AUC | 0.2993 | 917 | Median |
| OP | 2 | 0.5215 | 140 | Median | 0.4588 | -764 | Median | 0.3405 | 268 | Median | 0.8443 | -904 | Median | 0.0961 | 128 | Median | 0.0961 | 1032 | Median |
| ADRFI | 2 | 0.7909 | 163 | Median | 0.7909 | -736 | Median | 0.7909 | 163 | Median | 0.7909 | -899 | Median | 0.8069 | 84 | AUC | 0.7909 | 899 | Median |
| ENT |  |  |  |  |  |  |  |  |  |  |  |  |  |  |  |  |  |  |  |
| NP | 1 | 0.7667 | -167 | Median | 0.7035 | -389 | Median | 0.7035 | -310 | Median | 0.7035 | -222 | Median | 0.7667 | -144 | Median | 0.7667 | 78 | Median |
| OP | 1 | 0.7613 | -83 | Median | 0.7613 | -297 | Median | 0.7613 | -207 | Median | 0.7613 | -214 | Median | 0.7613 | -124 | Median | 0.7613 | 90 | Median |
| ADRFI | 1 | 0.5433 | 213 | Median | 0.0856 | -236 | Median | 0.0013 | -506 | Median | 0.02 | -448 | Median | 1e-04 | -718 | Median | 0.0856 | -270 | Median |
| NP | 2 | 0.4042 | 19 | Median | 0.9545 | -168 | Median | 2e-04 | -320 | Median | 0.4042 | -188 | Median | 0 | -340 | Median | 4e-04 | -152 | Median |
| OP | 2 | 0.1043 | -138 | Median | 0.8464 | 30 | Median | 0 | -344 | Median | 0.1603 | 168 | Median | 0.0015 | -206 | Median | 0 | -374 | Median |
| ADRFI | 2 | 0.4989 | 161 | Median | 0.2855 | -178 | Median | 0.0295 | -146 | Median | 0.0977 | -339 | Median | 0.005 | -307 | Median | 0.2855 | 32 | Median |

Detailed discriminative power results for HEGP patient groups using logistic regression (1/3)

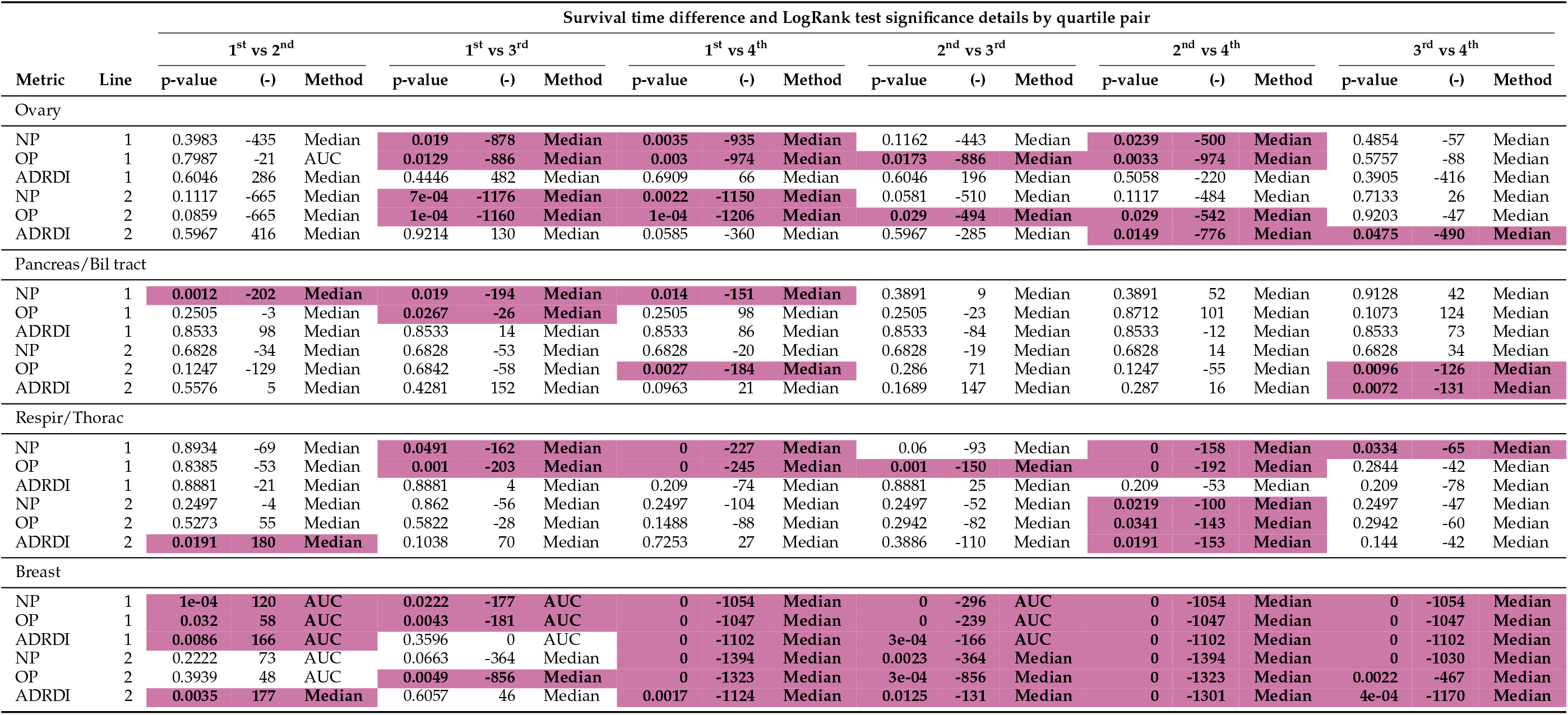

Detailed discriminative power results for HEGP patient groups using logistic regression (1 / 3)
| Survival time difference and LogRank test significance details by quartile pair |  |  |  |  |  |  |  |  |  |  |  |  |  |  |  |  |  |  |  |
| --- | --- | --- | --- | --- | --- | --- | --- | --- | --- | --- | --- | --- | --- | --- | --- | --- | --- | --- | --- |
| Metric | Line | 1<sup>st</sup> vs 2<sup>nd</sup> |  |  | 1<sup>st</sup> vs 3<sup>rd</sup> |  |  | 1<sup>st</sup> vs 4<sup>th</sup> |  |  | 2<sup>nd</sup> vs 3<sup>rd</sup> |  |  | 2<sup>nd</sup> vs 4<sup>th</sup> |  |  | 3<sup>rd</sup> vs 4<sup>th</sup> |  |  |
|  |  | p-value | (-) | Method | p-value | (-) | Method | p-value | (-) | Method | p-value | (-) | Method | p-value | (-) | Method | p-value | (-) | Method |
| Ovary |  |  |  |  |  |  |  |  |  |  |  |  |  |  |  |  |  |  |  |
| NP | 1 | 0.3983 | -435 | Median | 0.019 | -878 | Median | 0.0035 | -935 | Median | 0.1162 | -443 | Median | 0.0239 | -500 | Median | 0.4854 | -57 | Median |
| OP | 1 | 0.7987 | -21 | AUC | 0.0129 | -886 | Median | 0.003 | -974 | Median | 0.0173 | -886 | Median | 0.0033 | -974 | Median | 0.5757 | -88 | Median |
| ADRD | 1 | 0.6046 | 286 | Median | 0.4446 | 482 | Median | 0.6909 | 66 | Median | 0.6046 | 196 | Median | 0.5058 | -220 | Median | 0.3905 | -416 | Median |
| NP | 2 | 0.1117 | -665 | Median | 7e-04 | -1176 | Median | 0.0022 | -1150 | Median | 0.0581 | -510 | Median | 0.1117 | -484 | Median | 0.7133 | 26 | Median |
| OP | 2 | 0.0859 | -665 | Median | 1e-04 | -1160 | Median | 1e-04 | -1206 | Median | 0.029 | -494 | Median | 0.029 | -542 | Median | 0.9203 | -47 | Median |
| ADRD | 2 | 0.5967 | 416 | Median | 0.9214 | 130 | Median | 0.0585 | -360 | Median | 0.5967 | -285 | Median | 0.0149 | -776 | Median | 0.0475 | -490 | Median |
| Pancreas/Bil tract |  |  |  |  |  |  |  |  |  |  |  |  |  |  |  |  |  |  |  |
| NP | 1 | 0.0012 | -202 | Median | 0.019 | -194 | Median | 0.014 | -151 | Median | 0.3891 | 9 | Median | 0.3891 | 52 | Median | 0.9128 | 42 | Median |
| OP | 1 | 0.2505 | -3 | Median | 0.0267 | -26 | Median | 0.2505 | 98 | Median | 0.2505 | -23 | Median | 0.8712 | 101 | Median | 0.1073 | 124 | Median |
| ADRD | 1 | 0.8533 | 98 | Median | 0.8533 | 14 | Median | 0.8533 | 86 | Median | 0.8533 | -84 | Median | 0.8533 | -12 | Median | 0.8533 | 73 | Median |
| NP | 2 | 0.6828 | -34 | Median | 0.6828 | -53 | Median | 0.6828 | -20 | Median | 0.6828 | -19 | Median | 0.6828 | 14 | Median | 0.6828 | 34 | Median |
| OP | 2 | 0.1247 | -129 | Median | 0.6842 | -58 | Median | 0.0027 | -184 | Median | 0.286 | 71 | Median | 0.1247 | -55 | Median | 0.0096 | -126 | Median |
| ADRD | 2 | 0.5576 | 5 | Median | 0.4281 | 152 | Median | 0.0963 | 21 | Median | 0.1689 | 147 | Median | 0.287 | 16 | Median | 0.0072 | -131 | Median |
| Respir/Thorac |  |  |  |  |  |  |  |  |  |  |  |  |  |  |  |  |  |  |  |
| NP | 1 | 0.8934 | -69 | Median | 0.0491 | -162 | Median | 0 | -227 | Median | 0.06 | -93 | Median | 0 | -158 | Median | 0.0334 | -65 | Median |
| OP | 1 | 0.8385 | -53 | Median | 0.001 | -203 | Median | 0 | -245 | Median | 0.001 | -150 | Median | 0 | -192 | Median | 0.2844 | -42 | Median |
| ADRD | 1 | 0.8881 | -21 | Median | 0.8881 | 4 | Median | 0.209 | -74 | Median | 0.8881 | 25 | Median | 0.209 | -53 | Median | 0.209 | -78 | Median |
| NP | 2 | 0.2497 | -4 | Median | 0.862 | -56 | Median | 0.2497 | -104 | Median | 0.2497 | -52 | Median | 0.0219 | -100 | Median | 0.2497 | -47 | Median |
| OP | 2 | 0.5273 | 55 | Median | 0.5822 | -28 | Median | 0.1488 | -88 | Median | 0.2942 | -82 | Median | 0.0341 | -143 | Median | 0.2942 | -60 | Median |
| ADRD | 2 | 0.0191 | 180 | Median | 0.1038 | 70 | Median | 0.7253 | 27 | Median | 0.3886 | -110 | Median | 0.0191 | -153 | Median | 0.144 | -42 | Median |
| Breast |  |  |  |  |  |  |  |  |  |  |  |  |  |  |  |  |  |  |  |
| NP | 1 | 1e-04 | 120 | AUC | 0.0222 | -177 | AUC | 0 | -1054 | Median | 0 | -296 | AUC | 0 | -1054 | Median | 0 | -1054 | Median |
| OP | 1 | 0.032 | 58 | AUC | 0.0043 | -181 | AUC | 0 | -1047 | Median | 0 | -239 | AUC | 0 | -1047 | Median | 0 | -1047 | Median |
| ADRD | 1 | 0.0086 | 166 | AUC | 0.3596 | 0 | AUC | 0 | -1102 | Median | 3e-04 | -166 | AUC | 0 | -1102 | Median | 0 | -1102 | Median |
| NP | 2 | 0.2222 | 73 | AUC | 0.0663 | -364 | Median | 0 | -1394 | Median | 0.0023 | -364 | Median | 0 | -1394 | Median | 0 | -1030 | Median |
| OP | 2 | 0.3939 | 48 | AUC | 0.0049 | -856 | Median | 0 | -1323 | Median | 3e-04 | -856 | Median | 0 | -1323 | Median | 0.0022 | -467 | Median |
| ADRD | 2 | 0.0035 | 177 | Median | 0.6057 | 46 | Median | 0.0017 | -1124 | Median | 0.0125 | -131 | Median | 0 | -1301 | Median | 4e-04 | -1170 | Median |

Detailed discriminative power results for HEGP patient groups using logistic regression (2/3)

| Survival time difference and LogRank test significance details by quartile pair |  |  |  |  |  |  |  |  |  |  |  |  |  |  |  |  |  |  |  |
| --- | --- | --- | --- | --- | --- | --- | --- | --- | --- | --- | --- | --- | --- | --- | --- | --- | --- | --- | --- |
| Metric | Line | 1<sup>st</sup> vs 2<sup>nd</sup> |  |  | 1<sup>st</sup> vs 3<sup>rd</sup> |  |  | 1<sup>st</sup> vs 4<sup>th</sup> |  |  | 2<sup>nd</sup> vs 3<sup>rd</sup> |  |  | 2<sup>nd</sup> vs 4<sup>th</sup> |  |  | 3<sup>rd</sup> vs 4<sup>th</sup> |  |  |
|  |  | p-value | (-) | Method | p-value | (-) | Method | p-value | (-) | Method | p-value | (-) | Method | p-value | (-) | Method | p-value | (-) | Method |
| Bladder/Uroth |  |  |  |  |  |  |  |  |  |  |  |  |  |  |  |  |  |  |  |
| NP | 1 | 0.1119 | -300 | Median | 0.8734 | 162 | Median | 0.0101 | -523 | Median | 0.1325 | 462 | Median | 0.2259 | -223 | Median | 0.0101 | -685 | Median |
| OP | 1 | 0.0177 | 937 | Median | 0.1043 | -276 | Median | 9e-04 | -527 | Median | 2e-04 | -1213 | Median | 0 | -1464 | Median | 0.0716 | -251 | Median |
| ADPDI | 1 | 0.707 | -64 | Median | 0.0061 | -492 | Median | 0.707 | 138 | Median | 0.0077 | -428 | Median | 0.9286 | 202 | Median | 0.0077 | 630 | Median |
| NP | 2 | 0.0363 | -370 | Median | 0.0363 | -344 | Median | 1e-04 | -576 | Median | 0.9938 | 26 | Median | 0.0363 | -206 | Median | 0.0363 | -232 | Median |
| OP | 2 | 0.1836 | -312 | Median | 0.0579 | -370 | Median | 0 | -604 | Median | 0.5296 | -58 | Median | 0.0012 | -291 | Median | 0.0034 | -233 | Median |
| ADPDI | 2 | 0.8227 | -124 | Median | 0.0432 | -142 | Median | 0.4786 | -218 | Median | 0.0474 | -19 | Median | 0.4786 | -94 | Median | 0.2391 | -75 | Median |

Detailed discriminative power results for HEGP patient groups using logistic regression (3/3)

## References

1. DeVita Jr VT and Chu E. A history of cancer chemotherapy. Cancer research 2008; 68:8643–53

2. Khongorzul P, Ling CJ, Khan FU, Ihsan AU, and Zhang J. Antibody–drug conjugates: a comprehensive review. Molecular Cancer Research 2020; 18:3–19

3. Davis LE, Shalin SC, and Tackett AJ. Current state of melanoma diagnosis and treatment. Cancer biology & therapy 2019; 20:1366–79

4. Organization WH et al. Adherence to long-term therapies: evidence for action. World Health Organization, 2003

5. Puts M, Tu H, Tourangeau A, et al. Factors influencing adherence to cancer treatment in older adults with cancer: a systematic review. Annals of Oncology 2014; 25:564–77

6. Karacin C, Bilgetekin I, B Basal F, and Oksuzoglu OB. How does COVID-19 fear and anxiety affect chemotherapy adherence in patients with cancer. Future Oncology 2020; 16:2283–93

7. Gray R, Bradley R, Braybrooke J, et al. Increasing the dose intensity of chemotherapy by more frequent administration or sequential scheduling: a patient-level meta-analysis of 37 298 women with early breast cancer in 26 randomised trials. The lancet 2019; 393:1440–52

8. Ratain MJ, Goldstein DA, and Lichter AS. Interventional pharmacoeconomics—a new discipline for a cost-constrained environment. JAMA oncology 2019; 5:1097–8

9. Walter J, Moeller C, Resuli B, et al. Guideline adherence of tumor board recommendations in lung cancer and transfer into clinical practice. Journal of Cancer Research and Clinical Oncology 2023; 149:11679–88

10. Hryniuk W. The importance of dose intensity in the outcome of chemotherapy. Important Adv Oncol 1988 :121–41

11. Denduluri N, Patt DA, Wang Y, et al. Dose delays, dose reductions, and relative dose intensit in patients with cancer who received adjuvant or neoadjuvant chemotherapy in community oncology practices. Journal of the National Comprehensive Cancer Network 2015; 13:1383–93

12. Nielson CM, Bylsma LC, Fryzek JP, Saad HA, and Crawford J. Relative dose intensity of chemotherapy and survival in patients with advanced stage solid tumor cancer: a systematic review and meta-analysis. The Oncologist 2021; 26:e1609–e1618

13. Longo DL, Duffey P, DeVita Jr V, Wesley M, Hubbard S, and Young R. The calculation of actual or received dose intensity: a comparison of published methods. J Clin Oncol 1991; 9:2042–51

14. Lyman GH. Impact of chemotherapy dose intensity on cancer patient outcomes. Journal of the National Comprehensive Cancer Network 2009; 7:99–108

15. Bylsma LC, Nielson CM, Fryzek JP, Saad HA, and Crawford J. Chemotherapy Relative Dose Intensity, Overall Survival, and Hematologic Toxicity in Solid-Tumor Cancer Patients: A Literature Review and Meta-Analysis. Blood 2020; 136:32–3

16. Wu VS, Elshami M, Stitzel HJ, et al. Why the treatment sequence matters: interplay between chemotherapy cycles received, cumulative dose intensity, and survival in resected early-stage pancreas cancer. Annals of surgery 2023; 278:e677–e684

17. Hoba J, Grancher A, Hautefeuille V, et al. Relative dose intensity of first-line triplet chemotherapy in metastatic colorectal cancer. Digestive and Liver Disease 2024

18. Gridelli C and Shepherd FA. Chemotherapy for elderly patients with non-small cell lung cancer: a review of the evidence. Chest 2005; 128:947–57

19. Santoleri F, Lasala R, Abrate P, et al. ADA ETA BIO2021: real-world evaluation of adherence, persistence, and cost-effectiveness of originator and biosimilar biologic drugs in the treatment of rheumatoid arthritis: a multicenter study in Italy. Current Medical Research and Opinio 2023; 39:1729–35

20. Zapletal E, Rodon N, Grabar N, and Degoulet P. Methodology of integration of a clinical data warehouse with a clinical information system: the HEGP case. MEDINFO 2010. IOS Press, 2010:193–7

21. Cossin S, Diouf S, Griffier R, Le Barrois d’Orgeval P, Diallo G, and Jouhet V. Linkage of Hospita Records and Death Certificates by a Search Engine and Machine Learning. JAMIA Open 2021 Mar; 4:ooab005. DOI: 10.1093/jamiaopen/ooab005

22. Rogier A, Rance B, and Coulet A. ChemoOnto, an ontology to qualify the course of chemotherapies. 2024 Jan. DOI:10.5281/zenodo.10548491

23. Jhee JH, Rogier A, Giraud D, et al. Representation and comparison of chemotherapy protocols with ChemoKG and graph embeddings. SWAT4HCLS. 2024

